# Comparative effectiveness of the bivalent BA.4-5 and BA.1 mRNA-booster vaccines in the Nordic countries

**DOI:** 10.1101/2023.01.19.23284764

**Authors:** Niklas Worm Andersson, Emilia Myrup Thiesson, Ulrike Baum, Nicklas Pihlström, Jostein Starrfelt, Kristýna Faksová, Eero Poukka, Hinta Meijerink, Rickard Ljung, Anders Hviid

## Abstract

**Background:** Data on the comparative vaccine effectiveness (CVE) of the bivalent mRNA-booster vaccines containing the original SARS-CoV-2 and omicron BA.4-5 and BA.1 subvariants are limited.

**Methods:** In a period of BA.4-5 subvariants predominance, we estimated the CVE of the bivalent Comirnaty (Pfizer-BioNTech) and Spikevax (Moderna) BA.4-5 and BA.1 mRNA-booster vaccines given as a fourth dose in Denmark, Finland, Norway, and Sweden. From 1 July 2022 to 12 December 2022, we conducted nationwide cohort analyses using target trial emulation to compare risks of Covid-19 hospitalization and death in four-dose (second booster) with three-dose (first booster) vaccinated and between four-dose vaccinated individuals.

**Results:** Compared with having received three vaccine doses, receipt of a bivalent BA.4-5 booster as a fourth dose was associated with a country-combined CVE against Covid-19 hospitalization of 80.5% (95% confidence interval, 69.5% to 91.5%). The corresponding CVE for bivalent BA.1 boosters was 74.0% (68.6% to 79.4%). CVE against Covid-19 death was 77.8% (48.3% to 100%) and 80.1% (72.0% to 88.2%) for bivalent BA.4-5 and BA.1 boosters as a fourth dose, respectively. The CVE of bivalent BA.4-5 vs. BA.1 boosters were 32.3% (10.6% to 53.9%) for Covid-19 hospitalization and 12.3% (−36.1% to 60.7%) for death (the latter estimable in Denmark only).

**Conclusions:** Vaccination with bivalent BA.4-5 or BA.1 mRNA-booster vaccines as a fourth dose was associated with increased protection against Covid-19 hospitalization and death during a period of BA.4-5 predominance. Bivalent BA.4-5 boosters conferred moderately greater vaccine effectiveness against Covid-19 hospitalization compared with bivalent BA.1 boosters.

## INTRODUCTION

Fourth dose (i.e., second booster) vaccination to improve protection against severe Covid-19 outcomes in target populations are now recommended in many countries. To combat the attenuated efficacy of the original monovalent BNT162b2 (Comirnaty, Pfizer-BioNTech) and mRNA-1273 (Spikevax, Moderna) mRNA Covid-19 vaccines observed against the omicron variants compared with other variants,^1–3^ bivalent mRNA-booster vaccines, containing spike sequences from the original (ancestral) SARS-CoV-2 strain and omicron subvariants (BA.4-5 or BA.1), were recently made available and implemented in booster vaccination programs including in the Nordic countries.

While some clinical studies have shown that the bivalent BA.4-5 and BA.1 mRNA-booster vaccines increases neutralizing antibody responses against omicron compared with the original monovalent mRNA Covid-19 vaccines, others have not.^4–8^ Data on the comparative effectiveness of the bivalent mRNA-booster vaccines as a fourth dose to protect against severe Covid-19 outcomes are scarce.^9–12^ Previous observational studies of fourth doses are primarily limited to the monovalent mRNA Covid-19 vaccines and mainly during periods prior to the emergence of the current predominating omicron subvariants BA.4 and BA.5.^13–29^ As such, data on the effectiveness of the bivalent mRNA-vaccines are needed to guide Covid-19 vaccination policy and to evaluate the benefit of developing variant-adapted Covid-19 vaccines.

In nationwide cohort analyses in Denmark, Finland, Norway, and Sweden, we assessed the comparative effectiveness of the bivalent BA.4-5 or BA.1 mRNA-booster vaccines received as the fourth dose against Covid-19 hospitalization and death during omicron subvariants BA.4-5 predominance.

## METHODS

### Data sources and source populations

All four Nordic countries hold nationwide demography- and healthcare registers with individual-level data that can be linked using the country-specific unique identifiers assigned to all residents. With linkage of these registers, we obtained information on Covid-19 vaccinations and laboratory-confirmed SARS-CoV-2 infections, hospitalizations and comorbidities, and demographic variables (e.g., age, sex, and vital status) (see Supplementary Tables S1-S2 for further details on utilized registers and definitions of variables). Within each country, we established a source population of individuals who were known residents and had received at least three vaccine doses (i.e., a primary two-dose vaccination course and one booster) with the AZD1222 (Vaxzevria, Oxford-AstraZeneca; as part of the primary vaccination only), BNT162b2, and/or mRNA-1273 vaccines between 27 December 2020 and 12 December 2022. See Supplementary Table S3 for description of ethical approvals/exempts.

### Study cohorts

To be included in our study, individuals were not allowed 1) to have received the third or fourth vaccine dose within 90 days after the last received vaccine dose (to ensure that the received third or fourth doses were truly first or second booster doses, respectively), 2) to be younger than a country-specific lower age limit of 50 years in Denmark, 60 in Finland, 65 in Norway, and 50 in Sweden, or 3) to have received the fourth dose before 1 September 2022 in Denmark, 18 July 2022 in Finland, and 1 July 2022 in Norway and Sweden (the two latter criteria were defined according to respective health authorities fourth dose rollout strategy for the general target population). The omicron subvariants BA.4-5 have been the predominating subvariants in all countries since these dates. When individuals received a fourth vaccine dose they were classified according to whether it was a bivalent BA.4-5, bivalent BA.1, or monovalent (original) mRNA-booster vaccine regardless of vaccine brand. We considered any comparison that included fourth dose vaccination with monovalent vaccines as an additional comparison analysis.

### Outcomes

Covid-19 hospitalization was defined as inpatient hospitalization with a registered Covid-19-related diagnosis and a positive PCR test for SARS-CoV-2 (within 14 days before and 2 days after the day of admission) (Supplementary Table S4 for country-specific definitions). We defined Covid-19 death as death within 30 days of a positive PCR test for SARS-CoV-2 in Denmark, Finland, and Sweden, whereas we used Covid-19-specific diagnoses registered as the main cause of death in Norway (owing to data availability).

### Comparisons

#### Fourth dose- compared with third dose-vaccinated

We assessed the effectiveness of receiving a bivalent BA.4-5 and BA.1 (or monovalent) mRNA-booster vaccine as the fourth dose by comparing with having received three monovalent vaccine doses only through a matched design. Individuals who received a fourth dose were matched on this day with individuals who had not yet received a fourth dose. Individuals were matched on age (5-year bins), calendar month when the third dose was received, and a propensity score including sex, region of residence, vaccination priority groups (i.e., individuals at high-risk of severe Covid-19 or healthcare workers), selected comorbidities, and previous history of SARS-CoV-2 infection (Supplementary Table S2). The day the fourth dose was administered within each matched pair served as the index date for both individuals. If individuals who were included as a matched three-dose vaccinated (i.e., a reference individual) received a fourth dose later than the assigned index date, they were allowed to potentially re-enter as a fourth-dose recipient in a new matched pair on that given date.

#### Fourth dose comparisons by type of vaccine

We compared the effectiveness of vaccination with the bivalent BA.4-5 and BA.1 (and monovalent) boosters as a fourth dose directly using inverse probability weights. The day of vaccination with the fourth dose served as the index date. Covariates included in the weights were: calendar month of fourth dose vaccination, age, sex, region of residence, vaccination priority groups, selected comorbidities, and previous history of SARS-CoV-2 infection (Supplementary Table S2).

### Statistical analysis

For the matched analyses, we used logistic regression to estimate the propensity score of receiving the fourth dose under study given covariates as predictors and matching was on age, calendar month, and with a caliper width of 0.01 on the propensity score. For the weighted analyses, we used logistic regression to calculate inverse probability weights as ((1−p_0_)/(1−p_c_))/(p_0_/p_c_); p_0_ being the crude probability of receiving a bivalent BA.4-5 booster (or a bivalent BA.1 booster if the comparison did not involve the BA.4-5 boosters) and p_c_ being the same probability given covariates.

We followed individuals from day 8 after the index date (to ensure full immunization among fourth dose recipients) up until the day of an outcome event, 60 days had passed since the index date (i.e., allowing up to 52 days of follow-up since day 8), death, emigration, or end of the study period, whichever occurred first. Additionally, we censored individuals with a positive PCR test for SARS-CoV-2 in our follow-up period after 14 and 30 days after the test (as a positive test was part of the outcome ascertainments) for the Covid-19 hospitalization and death outcome analyses, respectively. Moreover, we did not allow individuals with recent SARS-CoV-2 infection (≤12 weeks) before the index date (to avoid outcome misclassification). Similarly, for the Covid-19 hospitalization outcome analysis, we did not allow individuals with a Covid-19 hospitalization before the index date. For the matched analyses, we also right-censored matched pairs if the reference third dose-vaccinated individual received a fourth dose during follow-up.^30^ Cumulative incidences were estimated by the Kaplan-Meier estimator, and from these we calculated the relative (i.e., comparative vaccine effectiveness [CVE]; being 1 – risk ratio) and absolute risk differences at day 60. The corresponding 95% confidence interval (CI) of the effect estimate was calculated using the delta method. Upper 95% CIs for the CVE estimates were truncated at 100% if higher. Country-specific estimates were combined by random-effects meta-analyses implemented using the *mixmeta* package in R.

## RESULTS

### Study populations

The study cohorts comprised 3,368,697 fourth dose-vaccinated individuals across the four countries, of whom 1,290,999 (38%) had received a bivalent BA.4-5, 992,282 (30%) a bivalent BA.1, and 1,085,416 (32%) a monovalent mRNA-booster vaccine. Denmark contributed with a relatively larger sample of bivalent mRNA-booster vaccineés (a total of 1,676,138, 73% of all included bivalent booster vaccinated) than Finland (135,267, 6%), Norway (202,753, 9%), and Sweden (269,123, 12%) (Table 1, Supplementary Figures S1-S2, and Tables S5-S9). Slightly more than half of all individuals who had received a bivalent BA.4-5 or BA.1 booster vaccine within each country cohort were women with mean ages of 72 years, except in Sweden (approximately 60 years) and for the bivalent BA.4-5 booster vaccinated in Denmark (66 years).

**Table 1.** Baseline characteristics of recipients of a bivalent BA.4-5 or BA.1 mRNA-booster vaccine as a fourth vaccine dose across the four Nordic countries.^a^.

|  | Denmark |  | Finland |  | Norway |  | Sweden |  |
| --- | --- | --- | --- | --- | --- | --- | --- | --- |
| Bivalent booster received as fourth dose | BA.4-5 booster | BA.1 booster | BA.4-5 booster | BA.1 booster | BA.4-5 booster | BA.1 booster | BA.4-5 booster | BA.1 booster |
| Total individuals | 1105857 | 570281 | 86295 | 48972 | 71275 | 131478 | 27572 | 241551 |
| Mean age (SD) | 65.9 (10) | 72.6 (10.5) | 72.2 (7.5) | 71.6 (7.3) | 72.6 (6.5) | 72.1 (6) | 60.4 (7.5) | 60.9 (8) |
| Percentage females | 51.6% | 55.8% | 54.9% | 53.4% | 50.5% | 50.9% | 54.5% | 54% |
| Calendar period (min-max) | 16/09/22 - 04/12/22 | 12/09/22 - 04/12/22 | 04/08/22 - 04/12/22 | 23/08/22 - 04/12/22 | 13/09/22 - 03/12/22 | 06/09/22 - 03/12/22 | 07/07/22 - 15/10/22 | 16/08/22 - 15/10/22 |
| **Vaccination priority groups** |  |  |  |  |  |  |  |  |
| Severe Covid-19 risk group | 45641 (4.1%) | 54593 (9.6%) | 9161 (10.6%) | 5389 (11.0%) | 386 (0.5%) | 402 (0.3%) | 37 (0.1%) | 813 (0.3%) |
| Health care workers | 86611 (7.8%) | 27353 (4.8%) | 2689 (3.1%) | 1206 (2.5%) | 3181 (4.5%) | 5806 (4.4%) | 3509 (12.7%) | 29624 (12.3%) |
| **Comorbidities** |  |  |  |  |  |  |  |  |
| Autoimmune-related condition | 42841 (3.9%) | 25709 (4.5%) | 3520 (4.1%) | 2163 (4.4%) | 1928 (2.7%) | 3408 (2.6%) | 1276 (4.6%) | 11168 (4.6%) |
| Cancer | 49805 (4.5%) | 34843 (6.1%) | 8924 (10.3%) | 4719 (9.6%) | 2748 (3.9%) | 5027 (3.8%) | 1301 (4.7%) | 12073 (5.0%) |
| Chronic pulmonary disease | 33257 (3.0%) | 26036 (4.6%) | 1725 (2.0%) | 1006 (2.1%) | 7595 (10.7%) | 13713 (10.4%) | 844 (3.1%) | 8700 (3.6%) |
| Cardiovascular condition or diabetes | 92755 (8.4%) | 70633 (12.4%) | 24743 (28.7%) | 15042 (30.7%) | 20470 (28.7%) | 37071 (28.2%) | 3843 (13.9%) | 37393 (15.5%) |
| Renal disease | 11415 (1.0%) | 9503 (1.7%) | 1098 (1.3%) | 613 (1.3%) | 521 (0.7%) | 951 (0.7%) | 282 (1.0%) | 2839 (1.2%) |
| **Previous SARS-CoV-2 infection** |  |  |  |  |  |  |  |  |
| After third vaccine dose | 387483<br>(35.0%) | 170360<br>(29.9%) | 13577<br>(15.7%) | 5182<br>(10.6%) | 3206<br>(4.5%) | 5841<br>(4.4%) | 2278<br>(8.3%) | 19147<br>(7.9%) |
| Before third vaccine dose | 54131<br>(4.9%) | 21394<br>(3.8%) | 2588<br>(3.0%) | 837 (1.7%) | 786 (1.1%) | 965 (0.7%) | 3922<br>(14.2%) | 30607<br>(12.7%) |
| Omicron-infection | 373645<br>(33.8%) | 164166<br>(28.8%) | 13717<br>(15.9%) | 5241<br>(10.7%) | 2915<br>(4.1%) | 5255<br>(4.0%) | 2962<br>(10.7%) | 23839<br>(9.9%) |
| No (any) previous infection | 664243<br>(60.1%) | 378527<br>(66.4%) | 70130<br>(81.3%) | 42953<br>(87.7%) | 67283<br>(94.4%) | 124672<br>(94.8%) | 21372<br>(77.5%) | 191797<br>(79.4%) |

<sup>a</sup>Rows are numbers (percentages) unless otherwise stated. Variable definitions are shown in Supplementary Table S2.

The distribution of comorbidities and history of previous SARS-CoV-2 infection was relatively similar between individuals vaccinated with bivalent BA.4-5 and BA.1 booster within each country. Across countries, the proportion of individuals with a medical history of cardiovascular disease or diabetes in the overall cohorts was larger in Finland and Norway than in Denmark and Sweden (almost one-third and one-tenth, respectively).

### Effectiveness of a fourth dose compared with three vaccine doses

The cumulative incidences of Covid-19 hospitalization and death within 60 days of follow-up comparing four-dose with three-dose vaccinated individuals in each country were low (Figure 1 and Supplementary Figure S3). For example, the cumulative incidences of Covid-19 hospitalization did not exceed 0.16% for both three and four dose-vaccinated in any comparison. Receiving a fourth vaccine dose with a bivalent BA.4-5 booster was associated with a lower risk of Covid-19 hospitalization when compared with having received only three vaccine doses in all four countries; CVE ranged between 74.1% and 91.2% across countries with a combined CVE estimate of 80.5% (95% CI 69.5% to 91.5%) (Table 2 and Supplementary Table S10). A fourth dose with a bivalent BA.1 booster was similarly associated with lower risks of Covid-19 hospitalization corresponding to a CVE of 74.0% (95% CI 68.6% to 79.4%; CVEs ranged between 67.9% and 85.8% across countries). For Covid-19 death, the combined CVE was 77.8% (95% CI 48.3% to 100%) for bivalent BA.4-5 and 80.1% (95% CI 72.0% to 88.2%) for BA.1 boosters. A monovalent mRNA vaccine as a fourth dose also improved protection against Covid-19 hospitalization (CVE 64.9%, 95% CI 57.7% to 72.2%) and death (64.2%, 95% CI 53.3% to 75.1%) (Supplementary Figure S4 and Table S11).

**Figure 1.**
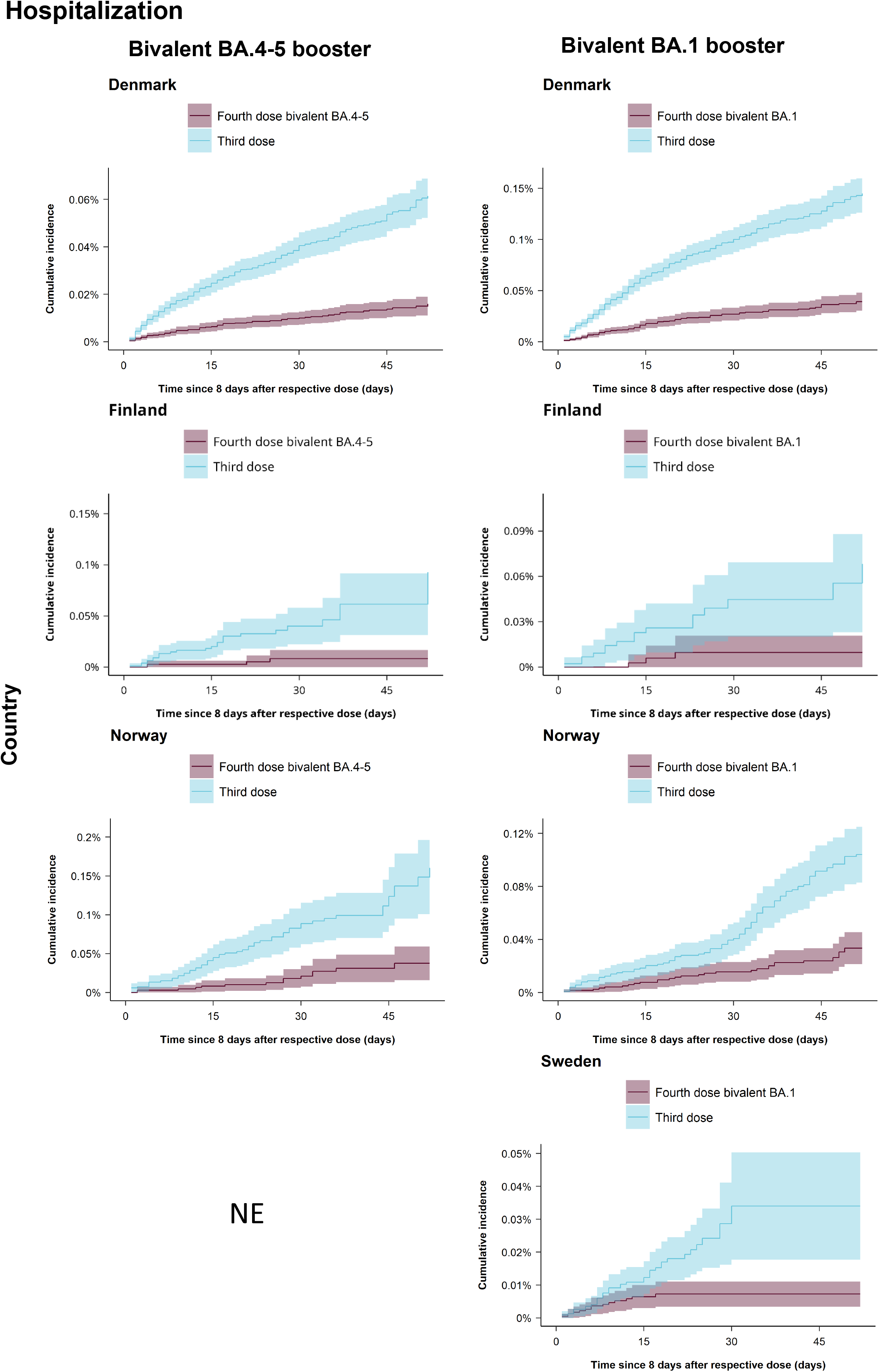
Cumulative incidence curves of Covid-19 hospitalization comparing individuals vaccinated with a bivalent BA.4-5 or BA.1 mRNA-booster vaccine as a fourth dose to individuals having received three vaccine doses only in each of the four Nordic countries. NE denotes not estimated meaning that the cumulative incidence curves could not be generated for this specific country (row) comparison (column). Cumulative incidence curves of Covid-19 death comparing individuals vaccinated with a bivalent mRNA-booster vaccine as a fourth dose to individuals having received three vaccine doses only in each of the four Nordic countries are presented in Supplementary Figure S3.

**Table 2.**
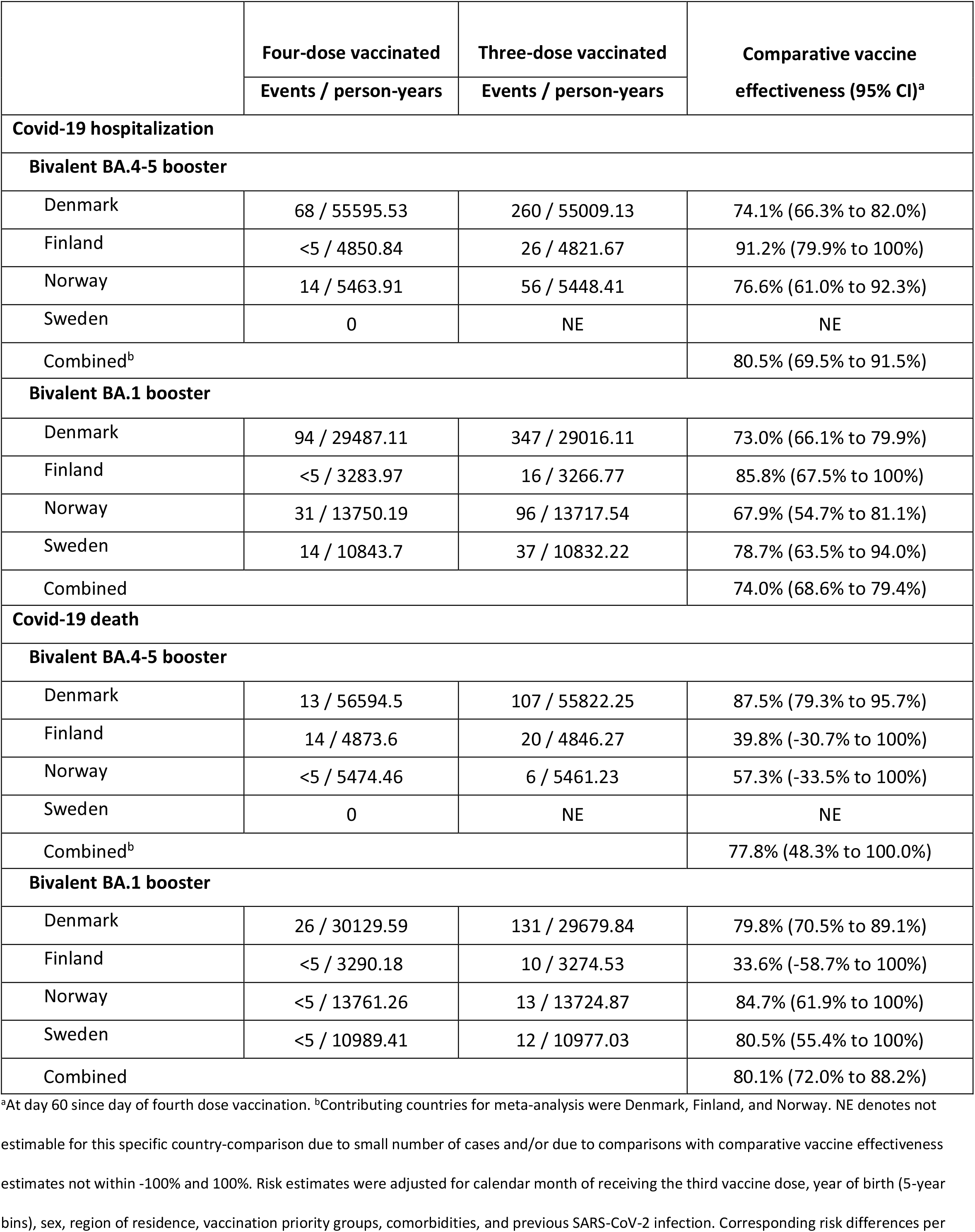
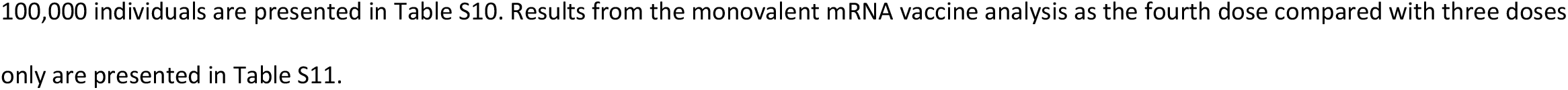
Risk of Covid-19 hospitalization and death comparing individuals vaccinated with a bivalent mRNA-booster vaccine received as a fourth dose to individuals vaccinated with only three doses in the four Nordic countries.

|  | Four-dose vaccinated | Three-dose vaccinated | Comparative vaccine effectiveness (95% CI)<sup>a</sup> |
| --- | --- | --- | --- |
|  | Events / person-years | Events / person-years |  |
| Covid-19 hospitalization |  |  |  |
| Bivalent BA.4-5 booster |  |  |  |
| Denmark | 68 / 55595.53 | 260 / 55009.13 | 74.1% (66.3% to 82.0%) |
| Finland | <5 / 4850.84 | 26 / 4821.67 | 91.2% (79.9% to 100%) |
| Norway | 14 / 5463.91 | 56 / 5448.41 | 76.6% (61.0% to 92.3%) |
| Sweden | 0 | NE | NE |
| Combined<sup>b</sup> |  |  | 80.5% (69.5% to 91.5%) |
| Bivalent BA.1 booster |  |  |  |
| Denmark | 94 / 29487.11 | 347 / 29016.11 | 73.0% (66.1% to 79.9%) |
| Finland | <5 / 3283.97 | 16 / 3266.77 | 85.8% (67.5% to 100%) |
| Norway | 31 / 13750.19 | 96 / 13717.54 | 67.9% (54.7% to 81.1%) |
| Sweden | 14 / 10843.7 | 37 / 10832.22 | 78.7% (63.5% to 94.0%) |
| Combined |  |  | 74.0% (68.6% to 79.4%) |
| Covid-19 death |  |  |  |
| Bivalent BA.4-5 booster |  |  |  |
| Denmark | 13 / 56594.5 | 107 / 55822.25 | 87.5% (79.3% to 95.7%) |
| Finland | 14 / 4873.6 | 20 / 4846.27 | 39.8% (-30.7% to 100%) |
| Norway | <5 / 5474.46 | 6 / 5461.23 | 57.3% (-33.5% to 100%) |
| Sweden | 0 | NE | NE |
| Combined<sup>b</sup> |  |  | 77.8% (48.3% to 100.0%) |
| Bivalent BA.1 booster |  |  |  |
| Denmark | 26 / 30129.59 | 131 / 29679.84 | 79.8% (70.5% to 89.1%) |
| Finland | <5 / 3290.18 | 10 / 3274.53 | 33.6% (-58.7% to 100%) |
| Norway | <5 / 13761.26 | 13 / 13724.87 | 84.7% (61.9% to 100%) |
| Sweden | <5 / 10989.41 | 12 / 10977.03 | 80.5% (55.4% to 100%) |
| Combined |  |  | 80.1% (72.0% to 88.2%) |

<sup>a</sup>At day 60 since day of fourth dose vaccination. <sup>b</sup>Contributing countries for meta-analysis were Denmark, Finland, and Norway. NE denotes not estimable for this specific country-comparison due to small number of cases and/or due to comparisons with comparative vaccine effectiveness estimates not within -100% and 100%. Risk estimates were adjusted for calendar month of receiving the third vaccine dose, year of birth (5-year bins), sex, region of residence, vaccination priority groups, comorbidities, and previous SARS-CoV-2 infection. Corresponding risk differences per

### Comparative effectiveness of the fourth vaccine dose received

Figure 2 presents the cumulative incidences of Covid-19 hospitalization and death comparing the bivalent BA.4-5 to the BA.1 boosters given as a fourth dose in each country. The combined estimate showed that receipt of a bivalent BA.4-5 booster was associated with a lower relative risk of Covid-19 hospitalization as compared with a BA.1 booster (CVE 32.3%, 95% CI 10.6% to 53.9%) (Table 3 and Supplementary Table S12). CVE estimates, however, were not uniform across countries, and the combined estimate was primarily driven by the larger-sized comparison in Denmark (CVE of 35.1%, 95% CI 12.6% to 57.7%). We observed no statistically significant difference in the risk of Covid-19 death when comparing bivalent BA.4-5 with BA.1 boosters in Denmark (CVE of 12.3%, 95% CI -36.1% to 60.7%; not estimable in other countries). The comparison of fourth dose vaccination with a bivalent BA.4-5 or BA.1 booster and a monovalent vaccine showed tendencies toward an increased protection associated with the bivalent boosters, but most results were not statistically significant and 95% CIs were very wide (Supplementary Figures S5-S6 and Table S13).

**Figure 2.**
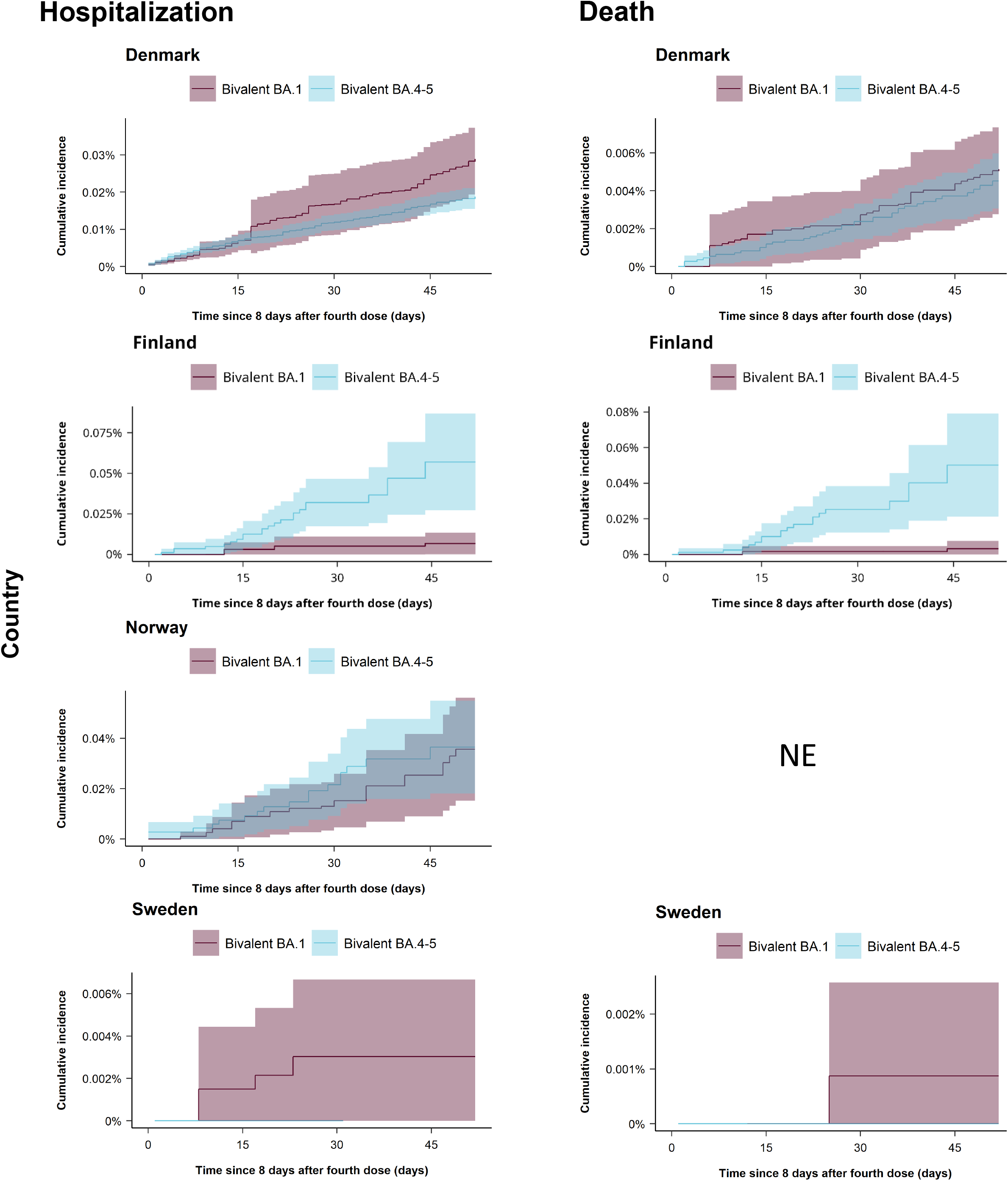
Cumulative incidence curves of Covid-19 hospitalization and death comparing individuals vaccinated with a bivalent BA.4-5 mRNA-booster vaccine received as a fourth dose to individuals vaccinated with a bivalent BA.1 mRNA-booster vaccine received as a fourth dose in each of the four Nordic countries. NE denotes not estimated meaning that the cumulative incidence curves could not be generated for this specific country (row) and outcome (column) comparison. Cumulative incidence curves comparing individuals vaccinated with a bivalent mRNA-booster vaccine received as a fourth dose to individuals vaccinated with a monovalent mRNA vaccine received as a fourth dose in each of the four Nordic countries are presented in Supplementary Figures S5-S6.

**Table 3.** Risk of Covid-19 hospitalization and death comparing individuals vaccinated with a bivalent BA.4-5 mRNA-booster vaccine received as a fourth dose to individuals vaccinated with a bivalent BA.1 mRNA-booster vaccine received as a fourth dose in the four Nordic countries.

|  | **Fourth dose bivalent**<br>**BA.4-5 booster** | **Fourth dose bivalent**<br>**BA.1 booster** | **Comparative vaccine**<br>**effectiveness (95% CI)<sup>a</sup>** |
| --- | --- | --- | --- |
|  | **Events / person-years** | **Events / person-years** |  |
| **Covid-19 hospitalization** |  |  |  |
| Denmark | 174 / 128630.69 | 278 / 78254.11 | 35.1% (12.6% to 57.7%) |
| Finland | 23 / 6001.33 | 6 / 3979.78 | NE |
| Norway | 17 / 6283.01 | 43 / 17593.63 | -2.3% (-80.5% to 75.9%) |
| Sweden | 0 / 350.14 | 15 / 12010.71 | NE |
| Combined<sup>b</sup> |  |  | 32.3% (10.6% to 53.9%) |
| **Covid-19 death** |  |  |  |
| Denmark | 40 / 129393.8 | 95 / 79063.99 | 12.3% (-36.1% to 60.7%) |
| Finland | 19 / 6036.34 | <5 / 3994.67 | NE |
| Norway | <5 / 6305.64 | <5 / 17651 | NE |
| Sweden | 0 / 353.92 | <5 / 12135.86 | NE |
| Combined |  |  | NA |

<sup>a</sup>At day 60 since day of fourth dose vaccination. <sup>b</sup>Contributing countries for meta-analysis were Denmark and Norway. NA denotes not applicable

## DISCUSSION

This study found that, during a period of omicron BA.4-5 subvariants predominance, both fourth dose vaccination (i.e., second booster) with the bivalent BA.4-5 or BA.1 mRNA-booster vaccines improved protection against Covid-19 hospitalization and death as compared with having received only three vaccine doses (i.e., a primary course and a first booster dose). Further, when the bivalent BA.4-5 and BA.1 mRNA-booster vaccines were directly compared, we observed that receiving a bivalent BA.4-5 booster as a fourth dose conferred moderately greater effectiveness against Covid-19 hospitalization than the BA.1 boosters.

Covid-19 vaccination policies recommending the bivalent mRNA-booster vaccines as a fourth dose are mainly supported by studies on immunogenicity where some have shown higher induction of antibody levels against omicron subvariants including BA.4-5 compared with monovalent boosters.^4–8^ Previous observational effectiveness studies of fourth dose monovalent vaccination, primarily on data from before BA.4-5 subvariant predominance,^13–29^ also lend some indirect support to these recommendations.

To date, little data exist on the effectiveness of the bivalent mRNA-boosters against Covid-19 outcomes. A recent US study found that when compared with unvaccinated, a fourth dose with a bivalent BA.4-5 booster was associated with moderate VEs (from 19% to 43%) against symptomatic infection.^31^ Two early reports from the Centers of Disease Control and Prevention, found moderate (CVE ranging from 38% to 45%)^10^ and high (73% to 83%)^9^ protection of the bivalent BA.4-5 boosters received after ≥2 monovalent mRNA vaccines against Covid-19 hospitalization when compared with past (≥2 months) monovalent mRNA vaccination only. These analyses, however, were not powered to examine the effect of the bivalent boosters received as a fourth dose. The two studies included only 783 and 79 individuals vaccinated with a bivalent booster, respectively, in test-negative case-control designs for the analysis of Covid-19 hospitalization (49 and 20 cases, respectively). Moreover, the generalizability of the results may be limited by selection bias;^9,10^ notably, over one half of bivalent booster recipients in the relatively larger-sized study^10^ had received four monovalent vaccine doses prior to the bivalent booster dose. In line with these findings, a UK report found that a bivalent BA.1 booster (i.e., as a ≥third dose) was associated with a CVE of 57% (95% CI 48 to 65%) against Covid-19 hospitalization in adults aged ≥50 years or those in clinical risk groups compared with ≥2 monovalent vaccine doses received at least 6 months prior.^11^ The CVE was estimated using a test-negative case-control design that included 176 cases and 621 controls (those testing positive and negative, respectively) that had received a bivalent booster (i.e., as a ≥third dose).^11^ Lastly, a recent Israeli pre-print, based on 85,313 bivalent BA.4-5 booster recipients as a ≥third dose, observed a hazard ratio of 0.19 (95% CI 0.08 to 0.42) for Covid-19 hospitalization (6 cases among bivalent booster recipients) compared with individuals having received ≥2 monovalent vaccines.^12^ However, the estimate for Covid-19 death was statistically insignificant (HR 0.14, 0.02 to 1.04; 1 case among bivalent booster recipients).^12^ Our results based on nationwide cohorts totaling 2,283,281 individuals vaccinated with bivalent boosters as a fourth dose strongly builds upon these early findings.

Our study has limitations. Although individuals were required to fulfill a restrictive set of pre-specified criteria in order to be considered Covid-19 hospitalized cases, we cannot exclude that our outcome definition captured cases where the infection with SARS-CoV-2 only partly contributed to or coincided with the timing of hospitalization. Similarly, our definition of Covid-19 death as any death occurring within 30 days of a positive SARS-CoV-2 PCR test used in Denmark, Finland, and Sweden was most likely subject to some outcome misclassification. However, our comparative population-based design would tend to mitigate larger differences in this potential outcome misclassification between compared groups as opposed to e.g., using unvaccinated individuals as a reference group. In the context of assessing the effectiveness of booster vaccinations, comparisons to unvaccinated individuals most likely hold greater risk of healthy vaccinee bias and such individuals would not reflect the targeted population for booster vaccination. For the outcome of Covid-19 death, we reassuringly observed no major differences in the effect estimates between Norway (where a Covid-19 cause-specific definition of death was used) and other countries.

Due to the nature of the healthcare registers in the four Nordic countries, we were able to consider potential confounders on an individual level, which were included by matching on propensity scores and by inverse probability weights, respectively. As a consequence of our study design, however, results from individual comparisons with different matched and weighted comparison groups should primarily be interpreted separately.

Our results likely have a high degree of generalizability to other similar populations. However, since we assessed the comparative effectiveness against Covid-19 hospitalization and death associated with the bivalent mRNA-booster vaccines given as a fourth dose, our results may only indirectly support any evaluation of the effectiveness of these vaccines within other Covid-19 vaccination schedule scenarios. Similarly, as per study design, we did not examine, and thus our results cannot directly help inform on, the vaccine effectiveness among individuals younger than 50 years old or other specific clinical subgroups that were not studied.

## CONCLUSION

As compared with individuals who have received three Covid-19 vaccine doses, vaccination with the bivalent BA.4-5 or BA.1 mRNA boosters as a fourth vaccine dose (i.e., second booster) increased protection against Covid-19 hospitalization and death. Fourth dose vaccination with bivalent BA.4-5 boosters conferred moderately greater protection against Covid-19 hospitalization than bivalent BA.1 boosters. Our findings provide much needed evidence on the effectiveness of bivalent boosters in the current pandemic context.

## Data Availability

No additional data available. Owing to data privacy regulations in each country, the raw data cannot be shared.

## Contributors

NA, ET, and AH conceptualized the study; NA drafted the manuscript; ET, UB, JS, and NP carried out the statistical analyses. All authors interpreted the results and critically reviewed the manuscript. AH supervised the study.

## Transparency

The lead author (the manuscript’s guarantor) affirms that this manuscript is an honest, accurate, and transparent account of the study being reported; that no important aspects of the study have been omitted; and that any discrepancies from the study as planned (and, if relevant, registered) have been explained.

## Ethics

The Danish study was performed as a surveillance study as part of the governmental institution Statens Serum Institut’s (SSI) advisory tasks for the Danish Ministry of Health. SSI’s purpose is to monitor and fight the spread of disease in accordance with section 222 of the Danish Health Act. According to Danish law, national surveillance activities conducted by SSI do not require approval from an ethics committee. It was approved by the Danish Governmental law firm and SSI’s compliance department that the study is fully compliant with all legal, ethical and IT-security requirements and there are no further approval procedures regarding such studies. For the Finnish study, by Finnish law, the Finnish Institute for Health and Welfare (THL) is the national expert institution to carry out surveillance on the impact of vaccinations in Finland (Communicable Diseases Act, https://www.finlex.fi/en/laki/kaannokset/2016/en20161227.pdf). Neither specific ethical approval (a waiver of ethical approval was received from Chief Doctor Otto Helve, Director of the Department for Health Security Finnish Institute for Health and Welfare) of this study nor informed consent from the participants was needed. The Norwegian study was approved by the Norwegian Regional Committee for Health Research Ethics South East (REK Sør-Øst A, ref 122745), and has conformed to the principles embodied in the Declaration of Helsinki. The emergency preparedness register was established according to the Health Preparedness Act §2-4. Consent to participate was not applicable as this is a register-based study. The Swedish study is approved by the Swedish Ethical Review Authority (2020-06859, 2021-02186) and has conformed to the principles embodied in the Declaration of Helsinki. Consent to participate is not applicable as this is a register-based study. Due to the nature of this research, there was no involvement of patients or members of the public in the design or reporting of this study.

## Disclaimer

This document expresses the opinion of the authors of the paper, and may not be understood or quoted as being made on behalf of, or reflecting the position of the European Medicines Agency or one of its Committees or Working Parties.

## Dissemination to participants and related patient and public communities

Studied participants were anonymised in the utilised data sources; and therefore, direct dissemination to study participants is not possible. The study results will be disseminated to the public and health professionals by a press release written using layman’s terms.

