## Supplementary for "Comparative effectiveness of the bivalent BA.4-5 and BA.1 mRNA-booster vaccines in the Nordic countries"

Supplementary Figure S1. Density plots of the distribution of age and index date for the matched comparisons of receiving a fourth vaccine dose vs three dose vaccinated only across the four countries. .. 15

**Supplementary Table S1. Description of utilized registers within each country.**

| Country/data source | Details |
|---|---|
| **Denmark** | |
| The Civil Registration System [1] | The register provides the unique personal identifier for all permanent residents of Denmark that allows linkage between all Danish health care registers and civil registrations systems. In addition, it holds general demographic information such as birthdate and sex as well as continuously updated information and dates on historical addresses, immigration and emigration status, and death. |
| The Danish Vaccination Register [2] | The register holds information on all vaccinations administered in Denmark including vaccination date, type/trade name, dose, and product batch number ever since Nov 15, 2015 (where reporting to the register became mandatory). Specifically related to this study, the Danish Health Agency have provided the governmentally assigned Covid-19 vaccine priority groups that were prioritized groups according to the risk of severe infection as well as whether being health and social care workers. |
| The Danish Microbiology Database [3] | Information on positive PCR tests for SARS-CoV-2 were retrieved from The Danish Microbiology Database (MiBa) that has data on all microbiology samples analysed at Danish microbiology departments as well as test results, date of sampling, date of analysis, type of test, and interpretation of test. The SARS-CoV-2 PCR tests are freely available to all individuals in Denmark regardless of symptoms status. |
| The National Patient Register [4] | The register holds information on all hospital contacts in Denmark including the duration of the contact, and diagnoses, which are assigned by the treating physician and registered according to ICD-10 classification system (since 1994). |
| **Finland** | |
| Finnish Population Information System [5] | The register is held by the Digital and Population Data Services Agency and contains personal data on all permanent residents in Finland such as the unique personal identifier, date of birth, place of residence, date of death, and date of immigration, and emigration. |
| Register of Social Assistance [6] | The register is held by the Finnish Institute for Health and Welfare and contains information on individuals receiving long-term care and/or social assistance (in e.g., nursing homes, people’s own homes or other institutions) including social rehabilitation. |
| Social and Healthcare Professionals Register [7] | The register holds data on individuals right to act as health care personnel. |
| National Vaccination Register [8] | The register is based on the Register of Primary Health Care Visits and contains information on all Covid-19 vaccinations administered in Finland including date of vaccination, batch number, and trade name. |
| National Infectious Diseases Register [9] | The register is held by the Finnish Institute for Health and Welfare and contains information on notifiable diseases in accordance with the Finnish Communicable Diseases Act that must be reported by the laboratories and the treating-physicians, or the physician performing an autopsy and hold information on sample dates of all laboratory-confirmed SARS-CoV-2 infections in Finland |

| Country/data source | Details |
|---|---|
| National Care Register for Health Care [10] | The register is held by the Finnish Institute for Health and Welfare and comprises information on all inpatient and outpatient hospital contacts in Finland, including admission and discharge dates, whether hospitalisation was planned or acute, codes for discharge diagnoses (according to ICD-10) and surgical procedures, and whether discharged as deceased, to own private residence or other health care facilities. |
| Special Reimbursement Register and Prescription Centre database | These databases are maintained by the Finnish Social Insurance Institution. The Special Reimbursement Register holds information on individuals entitled to special reimbursement for medical expenses. The Prescription Centre database holds information on individuals using selected medications of interest. |
| Register of Primary Health Care Visits [11] | The register is held by Finnish Institute for Health and Welfare and holds data on all primary health care services delivered in Finland. |
| **Norway** | |
| The Emergency Preparedness Register for Covid-19 [12] (consisting of the data sources below) | Data for the Norwegian analyses were collected through the Emergency preparedness register for Covid-19 ("Beredt C19"), which is administered by the Norwegian Institute of Public Health, according to the Norwegian Health Preparedness Act §2-4. The register was established in 2020 to provide authorities with up-to-date information on prevalence, causal relationships, and consequences of the Covid-19 epidemic in Norway and captures the entire population. The register includes information from the healthcare system and the national health registers presented below. |
| Norwegian Population Register | The register holds information on birthdate, immigration, emigration status, and death for all residents of Norway. |
| State register of employers and employees (NAV AA register) [13] | The register holds lists of all employment relationships in Norway for which employers and contractors are obliged to report to. Employees are classified according to the Norwegian Standard Classification of Occupations which we then used to identify whether individuals were health care personnel. |
| The Norwegian Information System for the Nursing and Care Sector (IPLOS) [14] | The register contains information on the health care services provided by municipalities and reporting of applicants and recipients of such services is mandatory in Norway. Available data includes information on home care services and out-of-hospital institutional care, including nursing home stays. |
| The Norwegian Immunisation Register (SYSVAK) [15] | The register holds information on administered vaccines in through the Norwegian vaccination programme, including date of administration and type of vaccine/trade name. For the Covid-19 vaccines, reporting to the register is mandatory. |
| Norwegian Surveillance System for Communicable Diseases (MSIS) | The register holds information on selected infectious diseases for which reporting to the register is mandatory, including all Covid-19 tests and testing date and results. |
| The Norwegian Patient Registry (NPR) [16] | The register holds data on all contacts with specialist health-care services in Norway, including admission and discharge dates diagnoses recorded according to ICD-10 during hospitalisation or outpatient contact. |

| Country/data source | Details |
|---|---|
| The Norwegian Intensive Care and Pandemic Registry (NIPaR) [17] | This is a national clinical registry that was expanded to include Covid-19 patients in conjunction with the Covid-19 pandemic. The register holds information on all patients who have tested positive for SARS-CoV-2 and were admitted to hospital including intensive care unit admissions. It is mandatory for all Norwegian hospitals to report to this register. |
| **Sweden** | |
| The Total Population Register [18] | The register is held by Statistics Sweden and contains data on the unique personal identifier assigned to all individuals in Sweden plus general demographic information such as date of birth, sex, country of birth, place of residence, and date of immigration and emigration. |
| The Cause of Death Register [19] | The register holds information on date of death and underlying as well as contributing causes of death. |
| The Longitudinal Integrated Database For Health Insurance And Labour Market Studies (LISA) [20] | The database is held by Statistics Sweden and holds many socioeconomic variables such as data on occupation which we used to identify whether individuals were health care personnel. |
| Register On Persons In Nursing Homes [21] | The register is held by the National Board of Health and Welfare and holds data on nursing care given in either nursing homes, own homes or other institutions to elderly and/or persons with physical, psychiatric or intellectual disabilities. |
| The National Vaccination Register [22] | The register is held by the Public Health Agency of Sweden and contains information on administered Covid-19 vaccines in Sweden including data on date of administration, the specific vaccine products, substance, formulation, batch number and dose number (for repeated doses). |
| Register On Surveillance Of Notifiable Communicable Diseases (Sminet) [23] | The register is held by the Public Health Agency of Sweden and contains information on notifiable diseases (for which reporting is mandatory) reported by either the analysis performing laboratories, the treating physician or autopsy performing physician, in accordance with the Swedish Communicable Diseases Act. Data included are e.g., date of disease occurrence, date of testing, date of positive test, and diagnoses. |
| The Swedish Patient Register [24,25] | The register is held by the National Board of Health and Welfare and comprises data on all in- and outpatient hospital specialist care in Sweden including data on dates of admission and discharge, whether hospitalisation was planned or acute, codes for discharge diagnoses (recorded according to ICD-10-SE) and surgical procedures, whether discharged as deceased, to own private residence or other health care facilities, and type of department. |

By use of nationwide register, we constructed country-specific cohorts with individual-level information on Covid-19 vaccination, endpoint, and covariate variables (see Table S2 for definitions). This linkage of variables and registers was made available by the unique personal identifier that is assigned to all residents within the respective Nordic country at either birth or immigration. Consequently, all utilized data were collected on the individual level. The health care systems in the Nordic countries are universal and tax-financed meaning the health care services are either freely available to all or subsidized so that all individuals pay only a fixed based minimum irrespective of the actual services provided and costs. We had full data availability for all variables during the study period and as reporting to national registers is mandatory/structurally implemented, this provides a near-complete follow-up of all residents over time.

**Supplementary Table S2. Definitions of included variables, details and data sources in each country.**

| Variable | Country | Data source and details | Values/codes |
|---|---|---|---|
| Age | Denmark | *The Civil Registration System.* Defined as age at the country-specific start date for the rollout of the fourth vaccine dose. | Categorical: 5-year bins |
| | Finland | *The Finnish Population Information System.* Defined as age at the country-specific start date for the rollout of the fourth vaccine dose. | |
| | Norway | *Norwegian Population Register.* Defined as age at the country-specific start date of the rollout for the fourth vaccine dose. | |
| | Sweden | *The Total Population Register.* Defined as age at the country-specific start date of the rollout for the fourth vaccine dose. | |
| Sex | Denmark | *The Civil Registration System.* Defined as registered sex. | Binary: male, female |
| | Finland | *The Finnish Population Information System.* Defined as registered sex. | |
| | Norway | *Norwegian Population Register.* Defined as registered sex. | |
| | Sweden | *The Total Population Register.* Defined as registered sex. | |
| Residency (citizenship) | Denmark | *The Civil Registration System.* Defined as known national resident. | Binary: yes/no |
| | Finland | Not available. | |
| | Norway | *Norwegian Population Register.* Defined as known national resident. | |
| | Sweden | *The Total Population Register.* Defined as known national resident. | |
| Calendar month | Denmark | *The Danish Vaccination Register.* Defined by the date where the respective vaccine dose examined was administered (i.e., third or fourth dose) and grouped into monthly intervals according to time since 27 December 2020. | Categorical (23 levels): calendar month 1 (27 December 2020 to 31 January 2021) to month 23 (1 November 2022 to 12 December 2022) |
| | Finland | *The National Vaccination Register.* Defined by the date where the respective vaccine dose examined was administered (i.e., third or fourth dose) and grouped into monthly intervals according to time since 27 December 2020. | |

| Variable | Country | Data source and details | Values/codes |
|---|---|---|---|
| | Norway | *The Norwegian Immunisation Register (SYSVAK).* Defined by the date where the respective vaccine dose examined was administered (i.e., third or fourth dose) and grouped into monthly intervals according to time since 27 December 2020. | |
| | Sweden[a] | *The National Vaccination Register.* Defined by the date where the respective vaccine dose examined was administered i.e., third or fourth dose) and grouped into monthly intervals according to time since 27 December 2020. | |
| Region of residency | Denmark | *The Civil Registration System.* Defined by last known address at the country-specific start date for the rollout of the fourth vaccine dose. | Categorical: Denmark, 5 levels; Finland, 5 levels; Norway, 5 levels; Sweden, 9 levels |
| | Finland | *The Finnish Population Information System.* Defined by last known municipality of residence. | |
| | Norway | *Norwegian Population Register.* Defined by last known address at the country-specific start date for the rollout of the fourth vaccine dose. | |
| | Sweden | *The Total Population Register.* Defined by last known address at the country-specific start date for the rollout of the fourth vaccine dose. | |
| Covid-19 vaccine priority groups[b] | Denmark | *The Danish Vaccination Register.* Defined as governmentally assigned Covid-19 vaccine priority groups, prioritized according to the risk of severe Covid-19 as well as whether being health and social care workers (last update 24 May 2021). | Categorical (3 levels): Severe Covid-19 risk group, healthcare personnel, others |
| | Finland | *Register of Social Assistance.* Severe Covid-19 risk group was defined as vulnerable individuals in 24-hours care (binary status per 27 December 2021).<br><br>*Social and Healthcare Professionals Register.* Healthcare personnel defined as individuals with the right to act as health care personnel as of 27 December 2021. | |
| | Norway | *The Norwegian Information System for the Nursing and Care Sector.* Severe Covid-19 risk group was defined as vulnerable individuals being residents at nursing homes (binary status per 27 December 2020). | |

| Variable | Country | Data source and details | Values/codes |
|---|---|---|---|
| | | *State register of employers and employees.* Healthcare personnel defined as binary status per 27 December 2020. | |
| | Sweden | *Register on persons in nursing homes.* Severe Covid-19 risk group was defined as vulnerable individuals being residents at nursing homes (binary status as of 31 December 2020)<br><br>*The Longitudinal integrated database for health insurance and labour market studies.* Healthcare personnel defined as healthcare worker occupation status as of 31 October 2018 (binary). | |
| Comorbidity 1: Chronic pulmonary disease | Denmark | *The National Patient Register.* Defined as primary diagnoses regardless of type of hospital contact registered before the start date for the country-specific rollout of the fourth vaccine dose (look-back 3 years). | Binary: yes/no<br><br>ICD-10 codes: J40-J47, J60–J67, J684, J701, J703, J841, J920, J961, J982, J983 |
| | Finland | *Care register for Health Care.* Defined as primary or secondary diagnoses before 27 December 2020 (look-back 6 years). | Binary: yes/no<br><br>ICD-10 codes: J41-J44, J47 |
| | Norway | *Norwegian Patient Register.* Defined as any recorded ICD-10 diagnosis during inpatient or outpatient contact in hospital or from private-practicing specialists and before first Covid-19 vaccination (look-back 3 years). | Binary: yes/no<br><br>ICD-10 codes: E84, J41-J47, J701, J703, J84, J98 |
| | Sweden | *National Patient Register.* Defined as any recorded ICD-10 diagnosis during inpatient or outpatient contact and before first Covid-19 vaccination (look-back 3 years). | Binary: yes/no<br><br>ICD-10 codes: E84, J41-J47 J84, J98 |
| Comorbidity 2: Cardiovascular conditions and diabetes | Denmark | *The National Patient Register.* Defined as primary diagnoses regardless of type of hospital contact registered before the start date for the country-specific rollout of the fourth vaccine dose (look-back 3 years). | Binary: yes/no<br><br>ICD-10 codes: E10-E11, I110, I130, I132, I20-I23, I420, I426-I429, I48, I500-I503, I508, I509 |
| | Finland | *Care register for Health Care, Register of Primary Health Care Visits, Special Reimbursement Register and Prescription Centre database.* Defined as primary or secondary diagnoses (look-back 6 years) or drug prescriptions (look-back 3 years) before 27 December 2020. | Binary: yes/no<br><br>ICD-10 codes: E10, E11, E13-E14, I11–I13, I15, I20–I25<br>ICPC-2 codes: T89, T90<br>ATC codes: A10A, A10B |

| Variable | Country | Data source and details | Values/codes |
|---|---|---|---|
| | Norway | *Norwegian Patient Register.* Defined as any recorded ICD-10 diagnosis during inpatient or outpatient contact in hospital or from private-practicing specialists and before first Covid-19 vaccination (look-back 3 years). | Binary: yes/no<br><br>ICD-10 codes: E10-E14, I05-I09, I110, I130, I132, I1420, I20-I23, I25-I28, I33-I39, I426-I429, I48, I50 |
| | Sweden | *National Patient Register.* Defined as any recorded ICD-10 diagnosis during inpatient or outpatient contact and before first Covid-19 vaccination (look-back 3 years).<br><br>*Swedish Prescribed Drug Register.* Antidiabetic drugs use defined as ≥2 filled prescriptions during 2020. | Binary: yes/no<br><br>ICD-10 codes: E10-E14, I05-I09, I110, I20-I28, I34-I37, I39, I42, I43, I46, I48-I50<br>ATC code: A10 |
| Comorbidity 3: Autoimmunity-related conditions[c] | Denmark | *The National Patient Register.* Defined as primary diagnoses regardless of type of hospital contact registered before the start date for the country-specific rollout of the fourth vaccine dose (look-back 3 years). | Binary: yes/no<br><br>ICD-10 codes: D510, D590, D591, D690, D693, D86, E050, E063, E271, E272, G122G, G35, G610, G700, I00, I01, K50, K51, K743, K900, L12, L40, L52, L80, L93, M05, M06, M08, M300, M313, M315, M316, M32, M33, M34, M35, M45 |
| | Finland | *Care register for Health Care, Special Reimbursement Register and Prescription Centre database.* Defined as primary or secondary diagnoses (look-back 6 years) or drug prescriptions (look-back 3 years) before 27 December 2020.<br><br>**Only if patient also used one of the listed drugs (marked with **)*<br>***Only if patient also had one of the diagnoses (marked with *)* | Binary: yes/no<br><br>ICD-10 codes: D7081, D7089, D80–D84, E250, E271, E272, E274, E310, E896, D86*, K50*, K51*, L40*, M02*, M05–M07*, M139*, M45*, M460*, M461*, M469*, M941*<br>ATC-codes**: H02AB02, H02AB04, H02AB06, H02AB07, L01BA01, L01XC02, L04AA06, L04AA10, L04AA13, L04AA18, L04AA24, L04AA26, L04AA29, L04AA33, L04AA37, L04AB, L04AC, L04AD01, L04AD02, L04AX01, L04AX03 |

| Variable | Country | Data source and details | Values/codes |
|---|---|---|---|
| | Norway | *Norwegian Patient Register.* Defined as any recorded ICD-10 diagnosis during inpatient or outpatient contact in hospital or from private-practicing specialists and before first Covid-19 vaccination (look-back 3 years). | Binary: yes/no<br><br>ICD-10 codes: G35, K50-K51, M05-M09, M13-M14 |
| | Sweden | *National Patient Register.* Defined as any recorded ICD-10 diagnosis during inpatient or outpatient contact and before first Covid-19 vaccination (look-back 3 years). | Binary: yes/no<br><br>ICD-10 codes: D86, G35, K50, K51, L40, M05-M09, M13, M14, M45 |
| Comorbidity 4: Cancer | Denmark | *The National Patient Register.* Defined as primary diagnoses regardless of type of hospital contact registered before the start date for the country-specific rollout of the fourth vaccine dose (look-back 3 years). | Binary: yes/no<br><br>ICD-10 codes: C00–C85 (without C44), C88, C90-C96 |
| | Finland | *Care register for Health Care and Special Reimbursement Register.* Defined as primary or secondary diagnoses before 27 December 2020 (look-back 6 years). | Binary: yes/no<br><br>ICD-10 codes: C00–C97 (without C44), D051, D39 |
| | Norway | *Norwegian Patient Register.* Defined as any recorded ICD-10 diagnosis during inpatient or outpatient contact in hospital or from private-practicing specialists and before first Covid-19 vaccination (look-back 3 years). | Binary: yes/no<br><br>ICD-10 codes: C00-C96 (without C44) |
| | Sweden | *National Patient Register.* Defined as any recorded ICD-10 diagnosis during inpatient or outpatient contact and before first Covid-19 vaccination (look-back 3 years). | Binary: yes/no<br><br>ICD-10 codes: C00-C96 (without C44), D45-D47 |
| Comorbidity 5: Moderate to severe renal disease | Denmark | *The National Patient Register.* Defined as primary diagnoses regardless of type of hospital contact registered before the start date for the country-specific rollout of the fourth vaccine dose (look-back 3 years). | Binary: yes/no<br><br>ICD-10 codes: I12, I13, N00–N05, N07, N11, N14, N17–N19, Q61 |
| | Finland | *Care register for Health Care.* Defined as primary or secondary diagnoses before 27 December 2020 (look-back 6 years). | Binary: yes/no<br><br>ICD-10 codes: I12, I13, N00–N05, N07, N08, N11, N14, N18, N19, E102, E112, E142 |
| | Norway | *Norwegian Patient Register.* Defined as any recorded ICD-10 diagnosis during inpatient or outpatient contact in hospital or from private- | Binary: yes/no |

| Variable | Country | Data source and details | Values/codes |
|---|---|---|---|
| | | practicing specialists and before first Covid-19 vaccination (look-back 3 years). | ICD-10 codes: I12-I13, N00-N05, N07, N11, N14, N17-N19, Q61 |
| | Sweden | *National Patient Register.* Defined as any recorded ICD-10 diagnosis during inpatient or outpatient contact and before first Covid-19 vaccination (look-back 3 years). | Binary: yes/no<br><br>ICD-10 codes: I12, I13, N00-N05, N07, N11, N14, N17-N19, Q61 |
| Any previous SARS-CoV-2 infection according to the date the third dose was received | Denmark | *The Danish Microbiology Database.* Defined as the date of any (last) registered positive PCR test for SARS-CoV-2 prior to the start date for the country-specific rollout of the fourth vaccine dose and according to the date of the third vaccine dose (those with a positive test during the last 12 weeks before the index date were excluded). | Categorical (3 levels): none, before, and after the third dose |
| | Finland | *National Infectious Diseases Register.* Defined as the date of any (last) registered positive PCR test for SARS-CoV-2 prior to the start date for the country-specific rollout of the fourth vaccine dose and according to the date of the third vaccine dose (those with a positive test during the last 12 weeks before the index date were excluded). | |
| | Norway | *Norwegian Surveillance System for Communicable Diseases (MSIS).* Defined as the date of any (last) registered positive PCR test for SARS CoV-2 prior to the start date for the country-specific rollout of the fourth vaccine dose and according to the date of the third vaccine dose (those with a positive test during the last 12 weeks before the index date were excluded). | |
| | Sweden | *Register on surveillance of notifiable communicable diseases (SmiNet).* Defined as the date of any (last) registered positive PCR test for SARS-CoV-2 prior the start date for the country-specific rollout of the fourth vaccine dose and according to the date of the third vaccine dose (those with a positive test during the last 12 weeks before the index date were excluded). | |
| Previous omicron infection | Denmark | *The Danish Microbiology Database.* Defined as the date of registered positive PCR test for SARS-CoV-2 between 28 December 2021[d] and the start date for the country-specific rollout of the fourth vaccine | Binary: yes/no |

| Variable | Country | Data source and details | Values/codes |
|---|---|---|---|
| | | dose (those with a positive test during the last 12 weeks before the index date were excluded). | |
| | Finland | *National Infectious Diseases Register.* Defined as the date of registered positive PCR test for SARS-CoV-2 between 1 January 2022[d] and the start date for the country-specific rollout of the fourth vaccine dose (those with a positive test during the last 12 weeks before the index date were excluded). | |
| | Norway | *Norwegian Surveillance System for Communicable Diseases (MSIS).* Defined as the date of registered positive PCR test for SARS CoV-2 between 28 December 2021[d] and the start date for the country-specific rollout of the fourth vaccine dose (those with a positive test during the last 12 weeks before the index date were excluded). | |
| | Sweden | *Register on surveillance of notifiable communicable diseases (SmiNet).* Defined as the date of registered positive PCR test for SARS-CoV-2 between 3 January 2022[d] and the start date for the country-specific rollout[d] of the fourth vaccine dose (those with a positive test during the last 12 weeks before the index date were excluded). | |

[a]Due to data availability in Sweden the country-specific end of study period was 23 October 2022. [b] To account for the risk of severe Covid-19, we adjusted for targeted Covid-19 high-risk groups of severe Covid-19, specifically established for each country. In Denmark, the Covid-19 vaccine priority groups were governmentally assigned and individuals were prioritized according to the risk of severe infection (identified by the treating physicians) as well as whether being health or social care workers. In the remaining countries, the variable was constructed based on the identification of vulnerable individuals (as defined by those receiving nursing care or living in nursing homes) and whether being health or social care workers. [c] Autoimmunity-related conditions includes a range disorders such as inflammatory bowel diseases, diseases involving the blood, immune mechanism or endocrine systems, inflammatory rheumatic diseases, psoriasis, lupus erythematosus, multiple sclerosis; subject to country-specific definitions. [d] The date defines the first day where omicron (sublineages BA.1 and BA.2) accounted for ≥90% of all registered SARS-CoV-2 infection cases within the country as per national surveillance data of SARS-CoV-2 variants.

**Supplementary Table S3. Description of ethical regulations within each country.**

| Country | Ethical Regulations |
|---|---|
| Denmark | The Danish analyses were performed as surveillance activities analyses as part of the advisory tasks of the governmental institution Statens Serum Institut (SSI) for the Danish Ministry of Health. SSI's purpose is to monitor and fight the spread of disease in accordance with section 222 of the Danish Health Act. According to Danish law, national surveillance activities conducted by SSI do not require approval from an ethics committee. Both the Danish Governmental law firm and the compliance department of SSI have approved that the study is fully compliant with all legal, ethical, and IT-security requirements and there are no further approval procedures required for such studies. |
| Finland | By Finnish law, the Finnish Institute for Health and Welfare (THL) is the national expert institution to carry out surveillance of the impact of vaccinations in Finland (Communicable Diseases Act, https://www.finlex.fi/en/laki/kaannokset/2016/en20161227.pdf). Neither specific ethical approval (a waiver of ethical approval was received from Chief Doctor Otto Helve, Director of the Department for Health Security Finnish Institute for Health and Welfare) of this study nor informed consent from the participants were needed. |
| Norway | The Norwegian analyses were conducted under the Norwegian Regional Committee for Health Research Ethics South East approval (REK Sør-Øst A) ref 122745 and conformed to the principles embodied in the Declaration of Helsinki. The utilized 'Emergency Preparedness Register for Covid-19' was established according to the Health Preparedness Act §2-4. Register-based studies (like this) in Norway are exempt from obtaining consent to participate. |
| Sweden | The Swedish analyses were conducted under the Swedish Ethical Review Authority approval 2020-06859, 2021-02186 and conformed to the principles embodied in the Declaration of Helsinki. Register-based studies (like this) in Sweden are exempt from obtaining consent to participate. |

**Supplementary Table S4. Covid-19 hospitalization and death outcome definitions.**

| Variable | Country | Data source and details |
|---|---|---|
| Covid-19 hospitalization | Denmark | *The National Patient Register and the Danish Microbiology Database.* Defined as a hospitalization on the day of, within 14 days of or in the two days after a PCR positive test for SARS-CoV-2, b) inpatient contact or at least 12 hours of contact, and c) a Covid-19 relevant diagnosis code (ICD-10: B342, B342A, B948A, B972, B972A, B972B, B972B1, Z038PA1) |
| | Finland | *National Care Register for Health Care and the National Infectious Diseases Register.* Defined as a hospitalization on the day of, within 14 days of or in the two days after a PCR positive test for SARS-CoV-2, b) inpatient hospital contact, and c) a Covid-19 relevant main diagnosis (ICD-10: J00-J22, J46, J80-J84, J851, J86, U071, U072). |
| | Norway | *The Norwegian Intensive Care and Pandemic Registry (NIPaR).* Defined as an individual with a positive PCR test for SARS-CoV-2 who were inpatient hospitalised and where Covid-19 was registered as the main cause of hospitalization. |
| | Sweden | *The Swedish Patient Register and the Register on surveillance of notifiable communicable diseases (SmiNet).* Defined as a hospitalization on the day of, within 14 days of or in the two days after a PCR positive test for SARS-CoV-2, b) inpatient contact or at least 12 hours of contact, and c) a Covid-19 relevant diagnosis code (ICD-10: U071, U072, U109) |
| Covid-19 death | Denmark | *The Civil Registration System and the Danish Microbiology Database.* Defined as (the date of) death within 30 days after PCR positive test for SARS-CoV-2. |
| | Finland | *The Finnish Population Information System and the National Infectious Diseases Register.* Defined as (the date of) death within 30 days after PCR positive test for SARS-CoV-2. |
| | Norway | *Norwegian Population Register and the Norwegian Surveillance System for Communicable Diseases (MSIS).* Defined as (the date of) death with a registered ICD-10 code of U071, U072, U109, or U099 as the main or contributing cause of death. |
| | Sweden | *The Total Population Register, the Cause of Death Register, and the Swedish Patient Register and the Register on surveillance of notifiable communicable diseases (SmiNet).* Defined as (the date of) death within 30 days after PCR positive test for SARS-CoV-2. |

**Supplementary Figure S1. Density plots of the distribution of age and index date for the matched comparisons of receiving a fourth vaccine dose vs three dose vaccinated only across the four countries.**

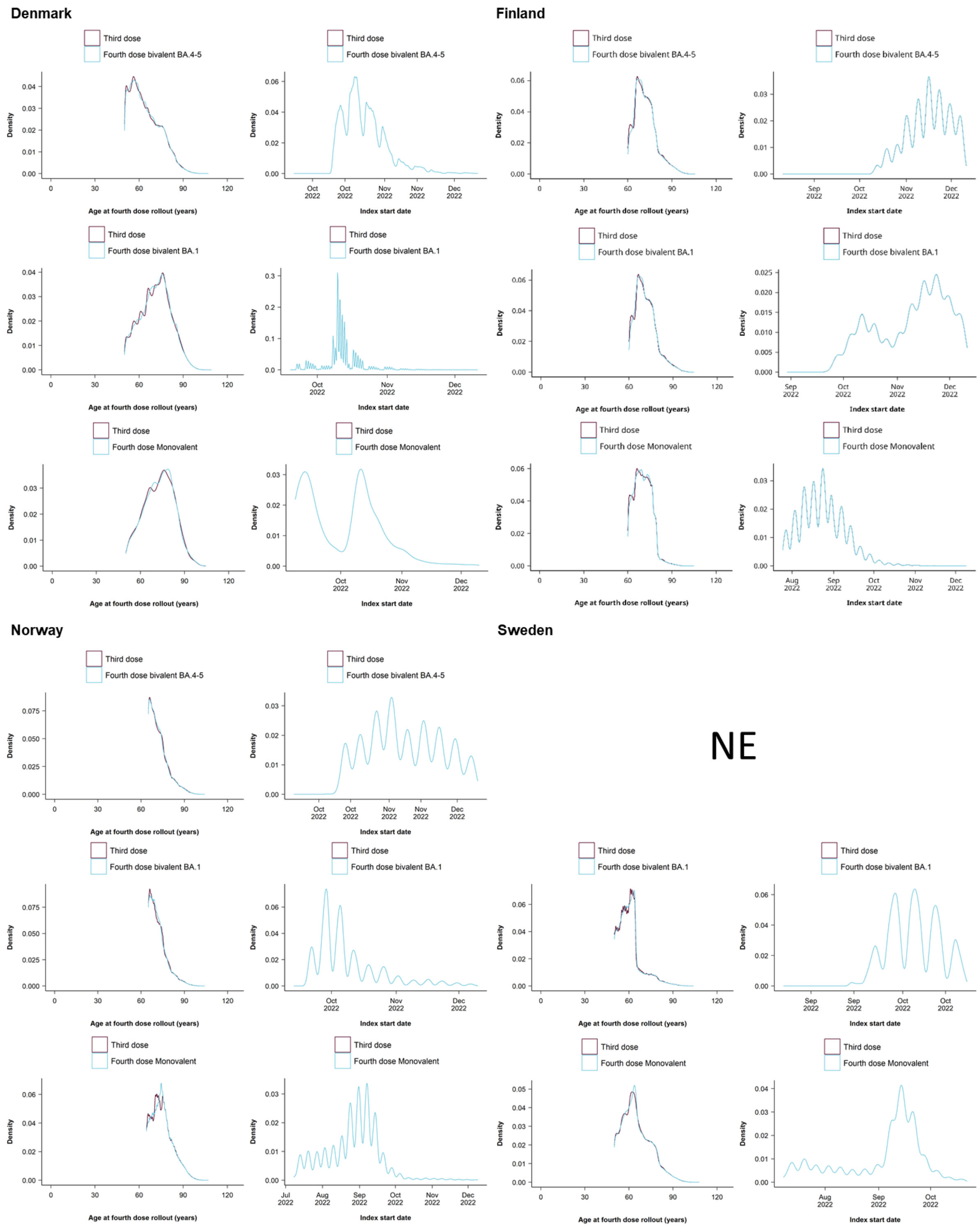

NE denotes not estimable (for the fourth dose bivalent BA.4-5 containing booster compared with three-dose vaccination in Sweden).

**Supplementary Figure S2. Density plots of the distribution of age and index date for the comparison of the different types of vaccines received as a fourth vaccine dose across the four countries.**

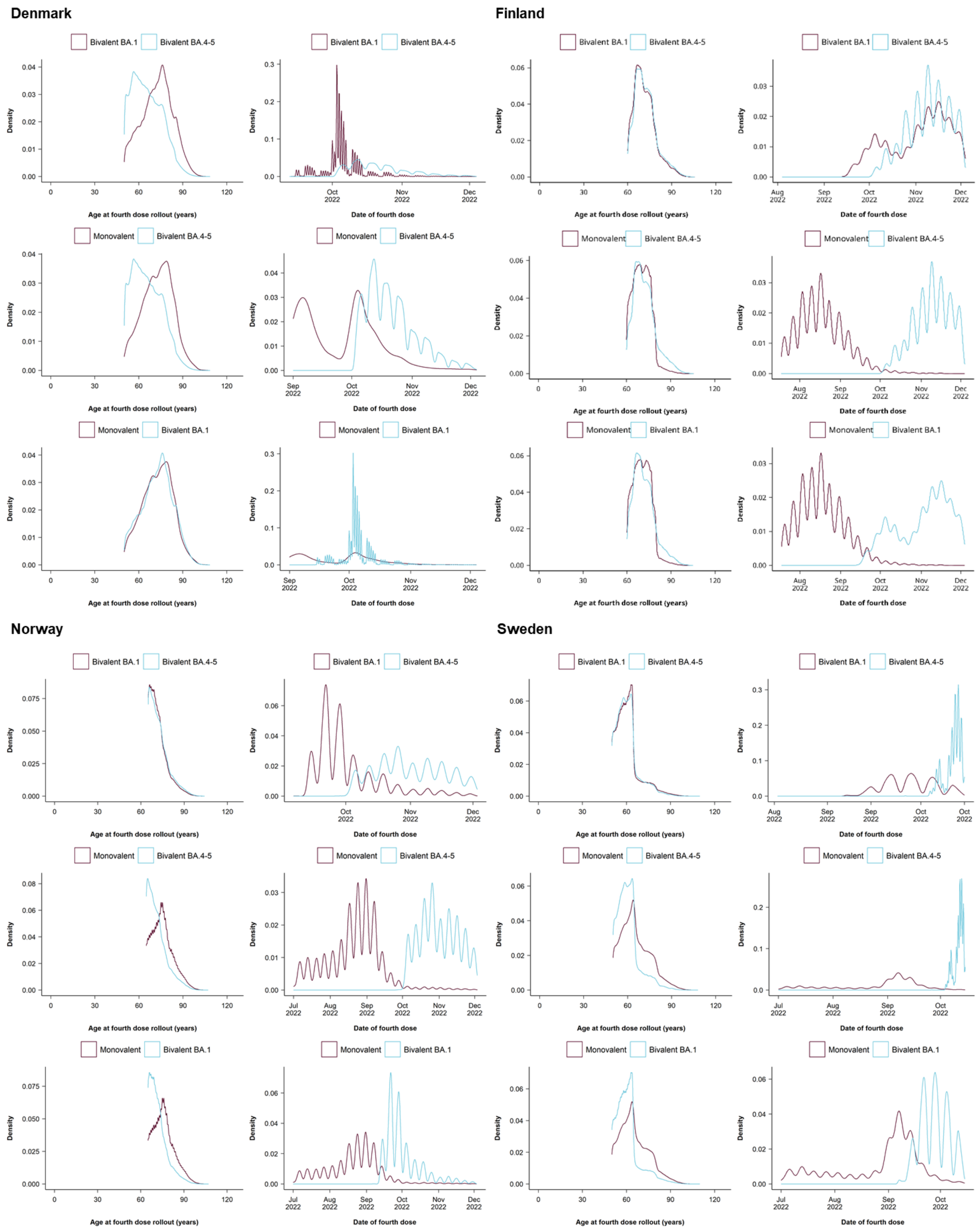

**Supplementary Table S5. Baseline characteristics of recipients of a monovalent mRNA vaccine as a fourth vaccine across the four Nordic countries.**

| | Denmark | Finland | Norway | Sweden |
|---|---|---|---|---|
| Total individuals | 4268 | 379275 | 428279 | 273594 |
| Mean age (SD) | 72.8 (10.2) | 70.4 (5.9) | 76 (7.0) | 65.4 (9.8) |
| Percentage females | 53.2% | 54.4% | 53.5% | 53.6% |
| Calendar period (min-max) | 01/09/22 - 03/12/22 | 18/07/22 - 02/12/22 | 01/07/22 - 02/12/22 | 01/07/22 - 15/10/22 |
| **Vaccination priority groups** | | | | |
| Severe Covid-19 risk group | 529 (12.4%) | 40879 (10.8%) | 12783 (3.0%) | 1936 (0.7%) |
| Health care workers | 208 (4.9%) | 740 (0.2%) | 10816 (2.5%) | 25185 (9.2%) |
| **Comorbidities** | | | | |
| Autoimmune-related condition | 270 (6.3%) | 13824 (3.6%) | 11394 (2.7%) | 15756 (5.8%) |
| Cancer | 395 (9.3%) | 36392 (9.6%) | 18620 (4.3%) | 19609 (7.2%) |
| Chronic pulmonary disease | 395 (9.3%) | 6454 (1.7%) | 48455 (11.3%) | 13542 (4.9%) |
| Cardiovascular condition or diabetes | 711 (16.7%) | 97487 (25.7%) | 142220 (33.2%) | 58377 (21.3%) |
| Renal disease | 114 (2.7%) | 3301 (0.9%) | 4679 (1.1%) | 5177 (1.9%) |
| **Previous SARS-CoV-2 infection** | | | | |
| After third vaccine dose | 1118 (26.2%) | 6367 (1.7%) | 18802 (4.4%) | 17743 (6.5%) |
| Before third vaccine dose | 198 (4.6%) | 1452 (0.4%) | 2187 (0.5%) | 29306 (10.7%) |
| Omicron-infection | 1076 (25.2%) | 5926 (1.6%) | 17027 (4.0%) | 21543 (7.9%) |
| No previous infection | 2952 (69.2%) | 371456 (97.9%) | 407290 (95.1%) | 226545 (82.8%) |

Rows are numbers (percentages) unless otherwise stated.

**Supplementary Table S6. Baseline characteristics of fourth dose and third dose vaccine recipients after matching in Denmark.**

| | Fourth dose bivalent BA.4-5 booster | Third dose | Fourth dose bivalent BA.1 booster | Third dose | Fourth dose monovalent booster[a] | Third dose[a] |
|---|---|---|---|---|---|---|
| Total individuals | 758369 | 758369 | 478343 | 478343 | 3950 | 3950 |
| Mean age (SD) | 64.6 (10.0) | 64.5 (10.1) | 71.4 (10.3) | 71.3 (10.4) | 72.6 (10.2) | 72.5 (10.3) |
| Percentage females | 51.2% | 50.6% | 55.5% | 53.8% | 53.4% | 52.5% |
| Calendar period (min-max) | 23/09/22 - 12/12/22 | 23/09/22 - 12/12/22 | 19/09/22 - 12/12/22 | 19/09/22 - 12/12/22 | 08/09/22 - 12/12/22 | 08/09/22 - 12/12/22 |
| **Vaccination priority groups** | | | | | | |
| Severe Covid-19 risk group | 27046 (3.6%) | 31170 (4.1%) | 34687 (7.3%) | 40368 (8.4%) | 474 (12.0%) | 386 (9.8%) |
| Health care workers | 57797 (7.6%) | 68451 (9.0%) | 25786 (5.4%) | 25193 (5.3%) | 196 (5.0%) | 299 (7.6%) |
| **Comorbidities** | | | | | | |
| Autoimmune-related condition | 28559 (3.8%) | 27873 (3.7%) | 21158 (4.4%) | 21486 (4.5%) | 253 (6.4%) | 186 (4.7%) |
| Cancer | 31852 (4.2%) | 31335 (4.1%) | 27403 (5.7%) | 30711 (6.4%) | 368 (9.3%) | 275 (7.0%) |
| Chronic pulmonary disease | 21692 (2.9%) | 21212 (2.8%) | 21128 (4.4%) | 19583 (4.1%) | 359 (9.1%) | 182 (4.6%) |
| Cardiovascular condition or diabetes | 59269 (7.8%) | 59805 (7.9%) | 56209 (11.8%) | 56934 (11.9%) | 646 (16.4%) | 474 (12.0%) |
| Renal disease | 7134 (0.9%) | 7731 (1.0%) | 7219 (1.5%) | 8220 (1.7%) | 107 (2.7%) | 95 (2.4%) |
| **Previous SARS-CoV-2 infection** | | | | | | |
| After third vaccine dose | 271711 (35.8%) | 243889 (32.2%) | 144297 (30.2%) | 130234 (27.2%) | 1026 (26.0%) | 1049 (26.6%) |
| Before third vaccine dose | 43231 (5.7%) | 36353 (4.8%) | 18921 (4.0%) | 15502 (3.2%) | 155 (3.9%) | 144 (3.6%) |
| Omicron-infection | 262087 (34.6%) | 235565 (31.1%) | 139081 (29.1%) | 125385 (26.2%) | 987 (25.0%) | 1013 (25.6%) |
| No previous infection | 443427 (58.5%) | 478127 (63.0%) | 315125 (65.9%) | 332607 (69.5%) | 2769 (70.1%) | 2757 (69.8%) |

[a]Presented matched cohort is for the analyses of Covid-19 hospitalization only. Rows are numbers (percentages) unless otherwise stated.

**Supplementary Table S7. Baseline characteristics of fourth dose and third dose vaccine recipients after matching in Finland.**

| | Fourth dose bivalent BA.4-5 booster | Third dose | Fourth dose bivalent BA.1 booster | Third dose | Fourth dose monovalent booster | Third dose |
|---|---|---|---|---|---|---|
| Total individuals | 81057 | 81057 | 46358 | 46358 | 356359 | 356359 |
| Mean age (SD) | 72 (7.3) | 72 (7.3) | 71.4 (7.1) | 71.4 (7.2) | 70.3 (5.9) | 70.2 (6) |
| Percentage females | 54.8% | 53.8% | 53.3% | 53.4% | 54.4% | 52.8% |
| Calendar period | 11/08/22 - 12/12/22 | 11/08/22 - 12/12/22 | 30/08/22 - 12/12/22 | 30/08/22 - 12/12/22 | 25/07/22 - 12/12/22 | 25/07/22 - 12/12/22 |
| **Vaccination priority groups** | | | | | | |
| Severe Covid-19 risk group | 8614 (10.6%) | 8169 (10.1%) | 5121 (11.0%) | 4760 (10.3%) | 38467 (10.8%) | 38938 (10.9%) |
| Health care workers | 2319 (2.9%) | 2057 (2.5%) | 1031 (2.2%) | 875 (1.9%) | 681 (0.2%) | 2731 (0.8%) |
| **Comorbidities** | | | | | | |
| Autoimmune-related condition | 3303 (4.1%) | 3106 (3.8%) | 2042 (4.4%) | 1824 (3.9%) | 12899 (3.6%) | 13071 (3.7%) |
| Cancer | 8352 (10.3%) | 7788 (9.6%) | 4439 (9.6%) | 4344 (9.4%) | 33889 (9.5%) | 32367 (9.1%) |
| Chronic pulmonary disease | 1589 (2.0%) | 1743 (2.2%) | 946 (2.0%) | 978 (2.1%) | 6033 (1.7%) | 6607 (1.9%) |
| Cardiovascular condition or diabetes | 22974 (28.3%) | 23409 (28.9%) | 14119 (30.5%) | 12893 (27.8%) | 91016 (25.5%) | 94048 (26.4%) |
| Renal disease | 1007 (1.2%) | 998 (1.2%) | 565 (1.2%) | 511 (1.1%) | 3094 (0.9%) | 3447 (1.0%) |
| **Previous SARS-CoV-2 infection** | | | | | | |
| After third vaccine dose | 12517 (15.4%) | 8346 (10.3%) | 4786 (10.3%) | 4580 (9.9%) | 5634 (1.6%) | 27107 (7.6%) |
| Before third vaccine dose | 2432 (3.0%) | 1324 (1.6%) | 790 (1.7%) | 705 (1.5%) | 1382 (0.4%) | 2451 (0.7%) |
| Omicron-infection | 12661 (15.6%) | 8406 (10.4%) | 4833 (10.4%) | 4607 (9.9%) | 5234 (1.5%) | 26496 (7.4%) |
| No previous infection | 66108 (81.6%) | 71387 (88.1%) | 40782 (88.0%) | 41073 (88.6%) | 349343 (98.0%) | 326801 (91.7%) |

Rows are numbers (percentages) unless otherwise stated.

**Supplementary Table S8. Baseline characteristics of fourth dose and third dose vaccine recipients after matching in Norway.**

| | **Fourth dose bivalent BA.4-5 booster** | **Third dose** | **Fourth dose bivalent BA.1 booster** | **Third dose** | **Fourth dose monovalent booster** | **Third dose** |
|---|---|---|---|---|---|---|
| Total individuals | 68338 | 68338 | 126593 | 126593 | 392699 | 392699 |
| Mean age (SD) | 72.4 (6.3) | 72.4 (6.4) | 72 (5.9) | 72 (6) | 75.7 (7) | 75.6 (7.1) |
| Percentage females | 50.5% | 51.6% | 50.9% | 51.2% | 53.4% | 53.1% |
| Calendar period | 20/09/22 - 12/12/22 | 20/09/22 - 12/12/22 | 13/09/22 - 12/12/22 | 13/09/22 - 12/12/22 | 08/07/22 - 12/12/22 | 08/07/22 - 12/12/22 |
| **Vaccination priority groups** | | | | | | |
| Severe Covid-19 risk group | 271 (0.4%) | 295 (0.4%) | 302 (0.2%) | 302 (0.2%) | 10531 (2.7%) | 8129 (2.1%) |
| Health care workers | 3089 (4.5%) | 3955 (5.8%) | 5650 (4.5%) | 7275 (5.7%) | 10455 (2.7%) | 10247 (2.6%) |
| **Comorbidities** | | | | | | |
| Autoimmune-related condition | 1818 (2.7%) | 1927 (2.8%) | 3240 (2.6%) | 3506 (2.8%) | 10306 (2.6%) | 10870 (2.8%) |
| Cancer | 2601 (3.8%) | 2435 (3.6%) | 4828 (3.8%) | 4550 (3.6%) | 16779 (4.3%) | 16644 (4.2%) |
| Chronic pulmonary disease | 7276 (10.6%) | 7205 (10.5%) | 13152 (10.4%) | 13315 (10.5%) | 43999 (11.2%) | 43809 (11.2%) |
| Cardiovascular condition or diabetes | 19435 (28.4%) | 18896 (27.7%) | 35451 (28.0%) | 34904 (27.6%) | 128233 (32.7%) | 130059 (33.1%) |
| Renal disease | 492 (0.7%) | 550 (0.8%) | 884 (0.7%) | 1011 (0.8%) | 4112 (1.0%) | 4354 (1.1%) |
| **Previous SARS-CoV-2 infection** | | | | | | |
| After third vaccine dose | 2912 (4.3%) | 2661 (3.9%) | 5396 (4.3%) | 5007 (4.0%) | 16694 (4.3%) | 14694 (3.7%) |
| Before third vaccine dose | 704 (1.0%) | 369 (0.5%) | 847 (0.7%) | 562 (0.4%) | 1743 (0.4%) | 1638 (0.4%) |
| Omicron-infection | 2639 (3.9%) | 2410 (3.5%) | 4840 (3.8%) | 4486 (3.5%) | 15113 (3.8%) | 13074 (3.3%) |
| No previous infection | 64722 (94.7%) | 65308 (95.6%) | 120350 (95.1%) | 121024 (95.6%) | 374262 (95.3%) | 376367 (95.8%) |

Rows are numbers (percentages) unless otherwise stated.

**Supplementary Table S9. Baseline characteristics of fourth dose and third dose vaccine recipients after matching in Sweden.[a]**

| | Fourth dose bivalent BA.1 booster | Third dose | Fourth dose monovalent booster | Third dose |
|---|---|---|---|---|
| Total individuals | 236462 | 236462 | 268610 | 268610 |
| Mean age (SD) | 60.8 (7.9) | 60.7 (8) | 65.3 (9.7) | 65.3 (9.7) |
| Percentage females | 53.9% | 51.6% | 53.5% | 52% |
| Calendar period | 23/08/22 - 23/10/22 | 23/08/22 - 23/10/22 | 08/07/22 - 23/10/22 | 08/07/22 - 23/10/22 |
| **Vaccination priority groups** | | | | |
| Severe Covid-19 risk group | 728 (0.3%) | 607 (0.3%) | 1685 (0.6%) | 1588 (0.6%) |
| Health care workers | 28736 (12.2%) | 31408 (13.3%) | 24688 (9.2%) | 29160 (10.9%) |
| **Comorbidities** | | | | |
| Autoimmune-related condition | 10912 (4.6%) | 10813 (4.6%) | 15400 (5.7%) | 14372 (5.4%) |
| Cancer | 11719 (5.0%) | 11049 (4.7%) | 19059 (7.1%) | 19543 (7.3%) |
| Chronic pulmonary disease | 8477 (3.6%) | 7858 (3.3%) | 13164 (4.9%) | 12664 (4.7%) |
| Cardiovascular condition or diabetes | 36370 (15.4%) | 35244 (14.9%) | 56879 (21.2%) | 59549 (22.2%) |
| Renal disease | 2746 (1.2%) | 2648 (1.1%) | 4973 (1.9%) | 5061 (1.9%) |
| **Previous SARS-CoV-2 infection** | | | | |
| After third vaccine dose | 18607 (7.9%) | 18902 (8.0%) | 17435 (6.5%) | 17484 (6.5%) |
| Before third vaccine dose | 30155 (12.8%) | 30196 (12.8%) | 28846 (10.7%) | 25702 (9.6%) |
| Omicron-infection | 23254 (9.8%) | 21821 (9.2%) | 21170 (7.9%) | 19635 (7.3%) |
| No previous infection | 187700 (79.4%) | 187364 (79.2%) | 222329 (82.8%) | 225424 (83.9%) |

[a]BA.4-5-containing vaccine vaccinated as a fourth dose vs three-dose vaccinated were not matched due to no Covid-19 outcome events among BA.4-5-containing vaccine vaccinated in Sweden. Rows are numbers (percentages) unless otherwise stated.

**Supplementary Figure S3. Cumulative incidence curves of Covid-19 death comparing individuals vaccinated with a bivalent BA.4-5 or BA.1 mRNA-booster vaccine as a fourth dose to individuals having received three vaccine doses only in each of the four Nordic countries.**

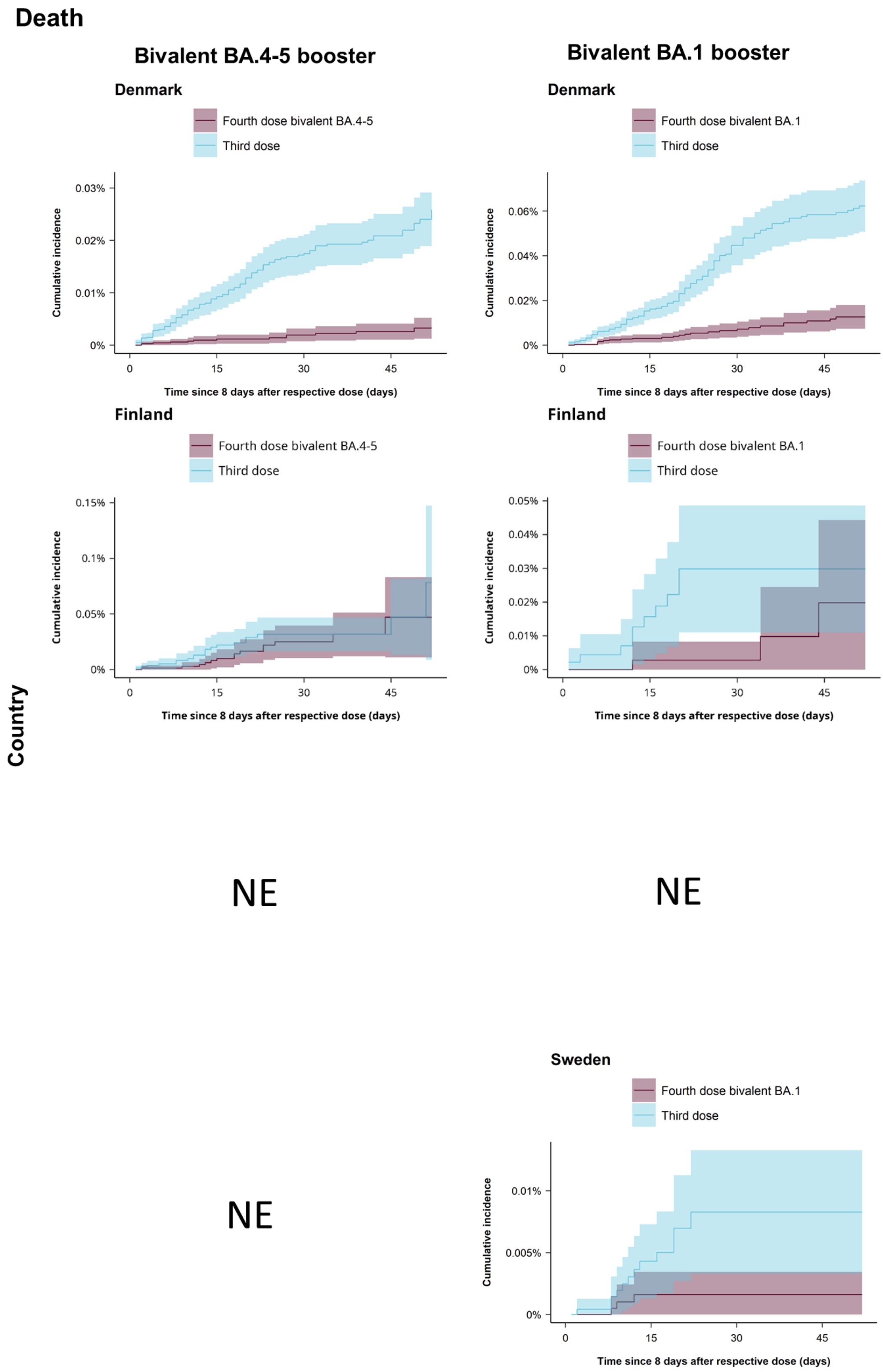

NE denotes not estimated meaning that the cumulative incidence curves could not be generated for this specific country (row) comparison (column).

**Supplementary Table S10. Risk differences for Covid-19 hospitalization and death comparing individuals vaccinated with a bivalent mRNA-booster vaccine received as a fourth dose to individuals vaccinated with only three doses in the four Nordic countries.**

| | Four-dose vaccinated | Three-dose vaccinated | Risk difference (95% CI) |
|---|---|---|---|
| | Events / person-years | Events / person-years | per 100,000 individuals[a] |
| **Covid-19 hospitalization** | | | |
| **Bivalent BA.4-5 booster** | | | |
| Denmark | 68 / 55595.53 | 260 / 55009.13 | -45.5 (-55.0 to -36.0) |
| Finland | <5 / 4850.84 | 26 / 4821.67 | -84.8 (-154.1 to -15.5) |
| Norway | 14 / 5463.91 | 56 / 5448.41 | -122.8 (-180.2 to -65.4) |
| Sweden | 0 | NE | NE |
| Combined[b] | | | -77.3 (-126.2 to -28.5) |
| **Bivalent BA.1 booster** | | | |
| Denmark | 94 / 29487.11 | 347 / 29016.11 | -105.8 (-125.1 to -86.6) |
| Finland | <5 / 3283.97 | 16 / 3266.77 | -58.6 (-101.1 to -16.1) |
| Norway | 31 / 13750.19 | 96 / 13717.54 | -70.6 (-94.8 to -46.3) |
| Sweden | 14 / 10843.7 | 37 / 10832.22 | -26.8 (-43.6 to -10.0) |
| Combined | | | -65.6 (-100.2 to -31.0) |
| **Covid-19 death** | | | |
| **Bivalent BA.4-5 booster** | | | |
| Denmark | 13 / 56594.5 | 107 / 55822.25 | -22.6 (-28.6 to -16.6) |
| Finland | 14 / 4873.6 | 20 / 4846.27 | -31.1 (-109.3 to 47.1) |
| Norway | <5 / 5474.46 | 6 / 5461.23 | -6.6 (-20.3 to 7.0) |
| Sweden | 0 | NE | NE |
| Combined[b] | | | -16.4 (-30.8 to -2.1) |
| **Bivalent BA.1 booster** | | | |
| Denmark | 26 / 30129.59 | 131 / 29679.84 | -49.7 (-62.3 to -37.0) |
| Finland | <5 / 3290.18 | 10 / 3274.53 | -10.0 (-40.9 to 20.9) |
| Norway | <5 / 13761.26 | 13 / 13724.87 | -12.7 (-21.5 to -3.8) |
| Sweden | <5 / 10989.41 | 12 / 10977.03 | -6.7 (-12.0 to -1.3) |
| Combined | | | -20.2 (-40.8 to 0.4) |

[a]At day 60 since day of fourth dose vaccination. [b]Contributing countries for meta-analysis were Denmark, Finland, and Norway. NE denotes not estimable for this specific country-comparison due to small number of cases and/or due to comparisons with comparative vaccine effectiveness estimates not within -100% and 100% (See Table 2). Risk estimates were adjusted for calendar month of receiving the fourth vaccine dose, year of birth (5-year bins), sex, region of residence, vaccination priority groups, comorbidities, and previous SARS-CoV-2 infection.

**Supplementary Figure S4. Cumulative incidence curves of Covid-19 hospitalization and death comparing individuals vaccinated with a monovalent mRNA vaccine as a fourth dose to individuals having received three vaccine doses only in each of the four Nordic countries.**

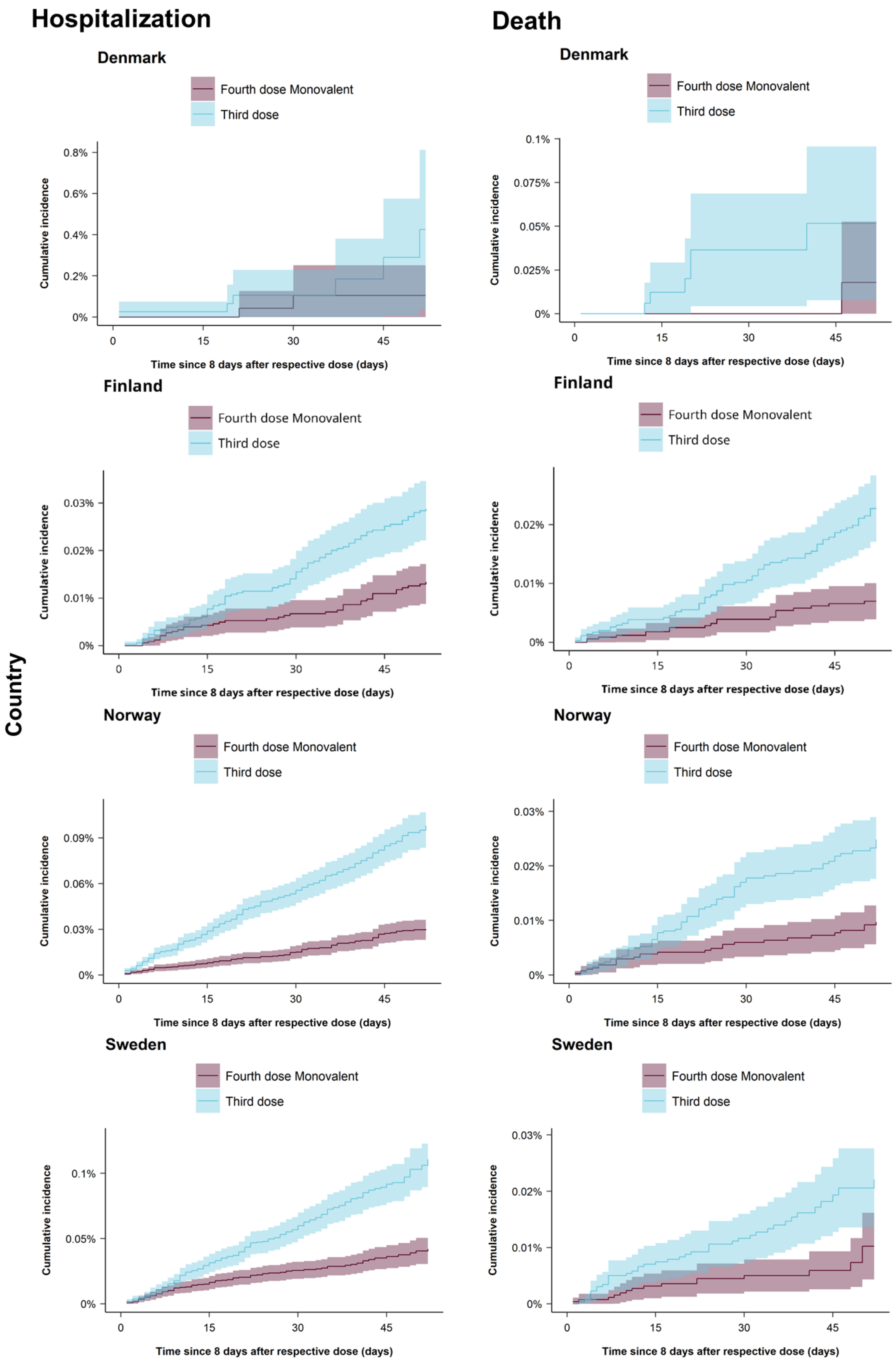

**Supplementary Table S11. Risk of Covid-19 hospitalization and death comparing individuals vaccinated with a monovalent mRNA vaccine received as a fourth dose to individuals vaccinated with only three doses in the four Nordic countries.**

| | Four-dose vaccinated | Three-dose vaccinated | Measure of association at day 60 since day of fourth dose vaccination | |
|---|---|---|---|---|
| | Events / person-years | Events / person-years | Risk difference (95% CI) per 100,000 individuals | Comparative vaccine effectiveness (95% CI) |
| **Covid-19 hospitalization** | | | | |
| Denmark | <5 / 295.01 | 6 / 293.65 | -320.6 (-737.3 to 96.1) | 75.5% (34.2% to 100%) |
| Finland | 38 / 40734.28 | 81 / 40434.48 | -15.4 (-23.1 to -7.8) | 53.6% (35.6% to 71.6%) |
| Norway | 83 / 39772.47 | 274 / 39636.58 | -68.0 (-81.6 to -54.4) | 69.3% (61.6% to 77.0%) |
| Sweden | 78 / 25772.51 | 194 / 25722.95 | -68.7 (-89.1 to -48.4) | 62.1% (50.9% to 73.2%) |
| Combined | | | -51.9 (-87.2 to -16.5) | 64.9% (57.7% to 72.2%) |
| **Covid-19 death** | | | | |
| Denmark | 0 | 0 | NE | NE |
| Finland | 20 / 40777.74 | 63 / 40471.5 | -15.8 (-22.2 to -9.3) | 69.3% (53.8% to 84.8%) |
| Norway | 28 / 39821 | 70 / 39669.66 | -15.1 (-22.1 to -8.1) | 60.9% (43.3% to 78.5%) |
| Sweden | 16 / 26203.12 | 40 / 26161.42 | -11.9 (-21.5 to -2.2) | 53.7% (22.5% to 84.9%) |
| Combined[a] | | | -14.8 (-19.0 to -10.5) | 64.2% (53.3% to 75.1%) |

[a]Contributing countries for meta-analysis were Finland, Norway, and Sweden. NE denotes not estimable for this specific country-comparison due to small number of cases and/or due to comparisons with comparative vaccine effectiveness estimates not within -100% and 100%. Risk estimates were adjusted for calendar month of receiving the third vaccine dose, year of birth (5-year bins), sex, region of residence, vaccination priority groups, comorbidities, and previous SARS-CoV-2 infection.

**Supplementary Table S12. Risk differences for Covid-19 hospitalization and death comparing individuals vaccinated with a bivalent BA.4-5 mRNA-booster vaccine received as a fourth dose to individuals vaccinated with a bivalent BA.1 mRNA-booster vaccine received as a fourth dose in the four Nordic countries.**

| | Fourth dose bivalent BA.4-5 booster | Fourth dose bivalent BA.1 booster | Risk difference (95% CI) per 100,000 individuals[a] |
|---|---|---|---|
| | Events / person-years | Events / person-years | |
| **Covid-19 hospitalization** | | | |
| Denmark | 174 / 128630.69 | 278 / 78254.11 | -10.2 (-19.6 to -0.7) |
| Finland | 23 / 6001.33 | 6 / 3979.78 | NE |
| Norway | 17 / 6283.01 | 43 / 17593.63 | 0.8 (-26.7 to 28.3) |
| Sweden | 0 / 350.14 | 15 / 12010.71 | NE |
| Combined[b] | | | -9.0 (-18.0 to -0.1) |
| **Covid-19 death** | | | |
| Denmark | 40 / 129393.8 | 95 / 79063.99 | -0.6 (-3.4 to 2.1) |
| Finland | 19 / 6036.34 | <5 / 3994.67 | NE |
| Norway | <5 / 6305.64 | <5 / 17651 | NE |
| Sweden | 0 / 353.92 | <5 / 12135.86 | NE |
| Combined | | | NA |

[a]At day 60 since day of fourth dose vaccination. [b]Contributing countries for meta-analysis were Denmark and Norway. NA denotes not applicable and NE not estimable for this specific country-comparison due to small number of cases and/or due to comparisons with comparative vaccine effectiveness estimates not within -100% and 100% (Table 3). Risk estimates were adjusted for calendar month of receiving the fourth vaccine dose, year of birth (5-year bins), sex, region of residence, vaccination priority groups, comorbidities, and previous SARS-CoV-2 infection.

**Supplementary Figure S5. Cumulative incidence curves of Covid-19 hospitalization comparing individuals vaccinated with a bivalent BA.4-5 and BA.1 mRNA-booster vaccine as a fourth dose to individuals vaccinated with a monovalent mRNA vaccine as a fourth dose in each of the four Nordic countries.**

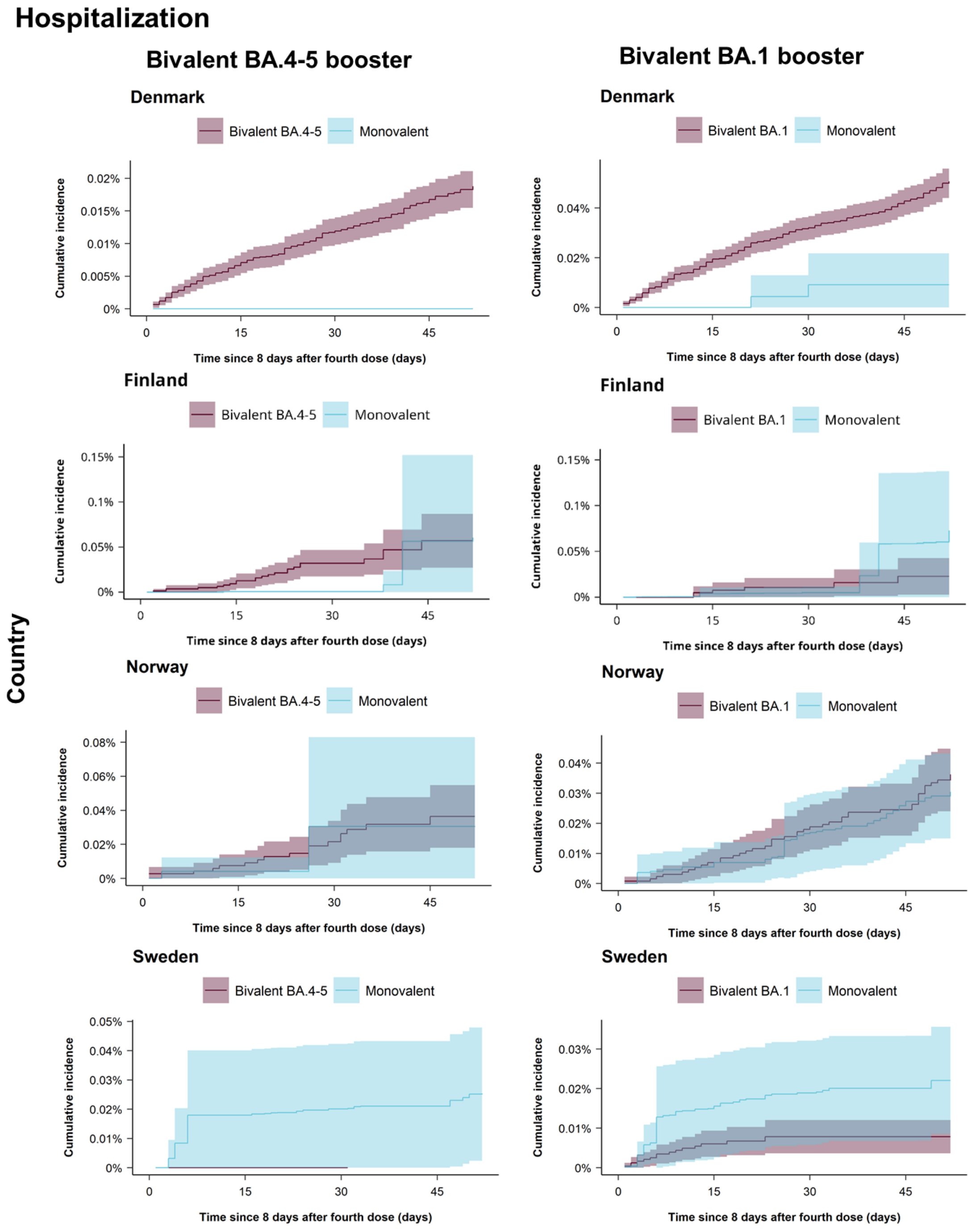

**Supplementary Figure S6. Cumulative incidence curves of Covid-19 death comparing individuals vaccinated with a bivalent BA.4-5 and BA.1 mRNA-booster vaccine as a fourth dose to individuals vaccinated with a monovalent mRNA vaccine as a fourth dose in each of the four Nordic countries.**

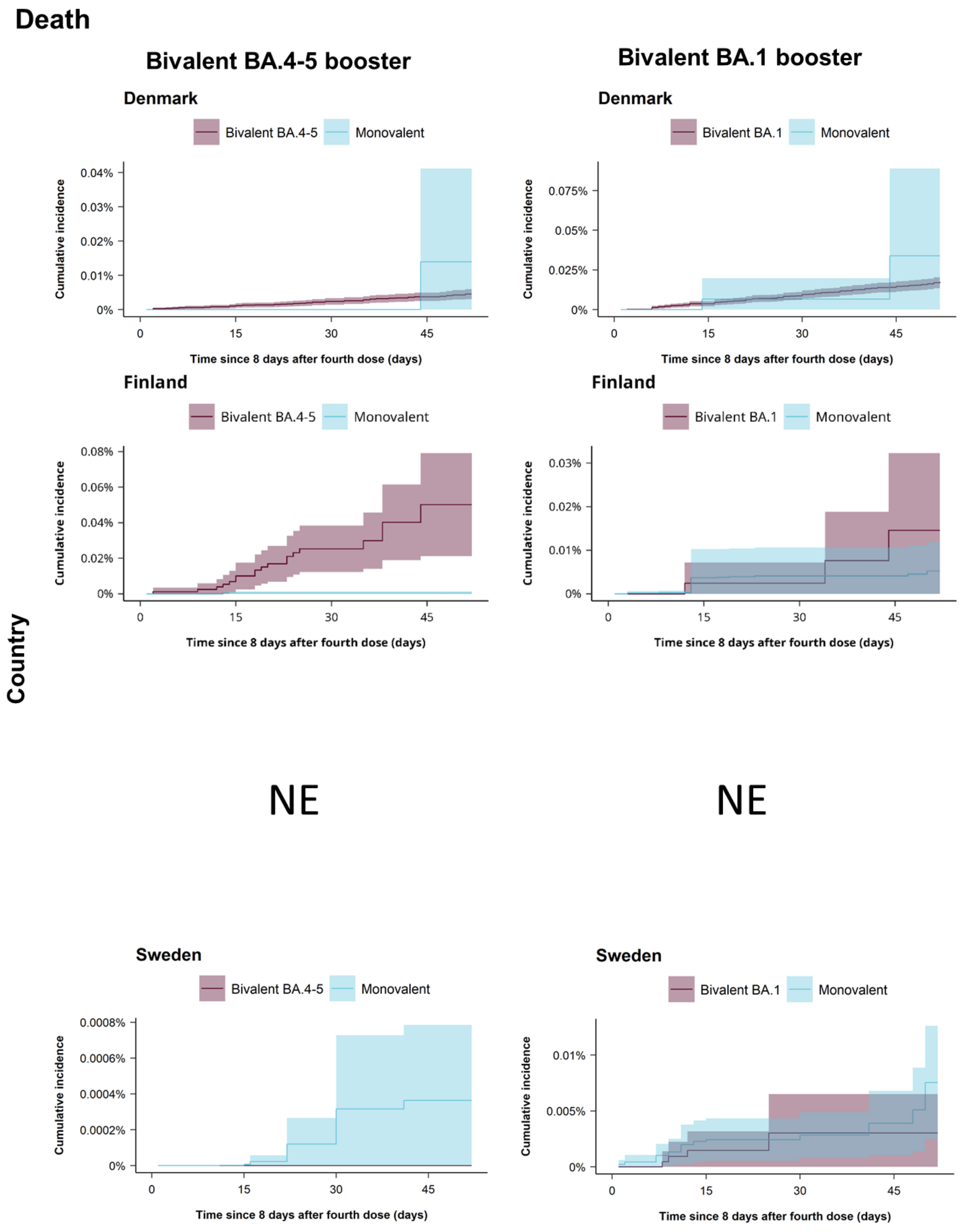

NE denotes not estimated meaning that the cumulative incidence curves could not be generated for this specific country (row; i.e., Norway) comparison (column).

**Supplementary Table S13. Risk of Covid-19 hospitalization and death comparing individuals vaccinated with a bivalent BA.4-5 and BA.1 mRNA-booster vaccine received as a fourth dose to individuals vaccinated with a monovalent mRNA vaccine received as a fourth dose in the four Nordic countries.**

| | Fourth dose bivalent booster | Fourth dose monovalent booster | Measure of association at day 60 since day of fourth dose vaccination | |
|---|---|---|---|---|
| | Events / person-years | Events / person-years | Risk difference (95% CI) per 100,000 individuals | Comparative vaccine effectiveness (95% CI) |
| **Covid-19 hospitalization** | | | | |
| **Bivalent BA.4-5 vs monovalent booster** | | | | |
| Denmark | 174 / 128630.69 | <5 / 571.07 | NE | NE |
| Finland | 23 / 6001.33 | 70 / 53779.95 | -3.4 (-104.0 to 97.2) | 5.6% (-152.4% to 100%) |
| Norway | 17 / 6283.01 | 142 / 60161.5 | 5.8 (-49.7 to 61.4) | -19.0% (-231.2% to 100%) |
| Sweden | 0 / 350.14 | 89 / 29974.02 | NE | NE |
| Combined[a] | | | 3.7 (-45.0 to 52.3) | -3.2% (-129.9% to 100%) |
| **Bivalent BA.1 vs monovalent booster** | | | | |
| Denmark | 278 / 78254.11 | <5 / 571.07 | NE | NE |
| Finland | 6 / 3979.78 | 70 / 53779.95 | -50.1 (-133.7 to 33.5) | 68.7% (24.4% to 100%) |
| Norway | 43 / 17593.63 | 142 / 60161.5 | 5.8 (-12.0 to 23.6) | -19.0% (-84.8% to 46.8%) |
| Sweden | 15 / 12010.71 | 89 / 29974.02 | -14.2 (-28.4 to -0.1) | 64.4% (35.5% to 93.3%) |
| Combined[b] | | | -7.1 (-25.9 to 11.6) | 45.0% (-2.7% to 92.6%) |
| **Covid-19 death** | | | | |
| **Bivalent BA.4-5 vs monovalent booster** | | | | |
| Denmark | 40 / 129393.8 | <5 / 581.31 | -9.4 (-36.7 to 17.9) | 67.5% (3.0% to 100%) |
| Finland | 19 / 6036.34 | 23 / 53818.89 | NE | NE |
| Norway | <5 / 6305.64 | 45 / 60357.63 | 3.0 (-14.6 to 20.6) | -51.9% (-429.1% to 100%) |
| Sweden | 0 / 353.92 | 18 / 30446.55 | NE | NE |
| Combined[c] | | | -0.6 (-15.4 to 14.2) | 64.1% (0.6% to 100.0%) |
| **Bivalent BA.1 vs monovalent booster** | | | | |
| Denmark | 95 / 79063.99 | <5 / 581.31 | -16.6 (-71.7 to 38.5) | 48.9% (-34.6% to 100%) |
| Finland | <5 / 3994.67 | 23 / 53818.89 | NE | NE |
| Norway | <5 / 17651 | 45 / 60357.63 | -2.7 (-9.6 to 4.3) | 52.6% (-27.5% to 100%) |
| Sweden | <5 / 12135.86 | 18 / 30446.55 | -4.5 (-10.7 to 1.6) | 59.8% (6.3% to 100%) |
| Combined[d] | | | -3.8 (-8.4 to 0.8) | 55.6% (16.4% to 94.9%) |

[a]Contributing countries for meta-analysis were Finland and Norway. [b]Contributing countries for meta-analysis were Finland, Norway, and Sweden. [c]Contributing countries for meta-analysis were Denmark and Norway. [d]Contributing countries for meta-analysis were Denmark, Norway, and Sweden. NE denotes not estimable for this specific country-comparison due to small number of cases and/or due to comparisons with comparative vaccine effectiveness estimates not within -100% and 100%. Risk estimates were adjusted for calendar month of receiving the fourth vaccine dose, year of birth (5-year bins), sex, region of residence, vaccination priority groups, comorbidities, and previous SARS-CoV-2 infection.

### Supplementary References.

1. Schmidt M, Pedersen L, Sørensen HT. The Danish Civil Registration System as a tool in epidemiology. Eur J Epidemiol 2014;29(8):541–9.

2. Krause TG, Jakobsen S, Haarh M, Mølbak K. The Danish vaccination register. Euro Surveill 2012;17(17):20155.

3. Voldstedlund M, Haarh M, Mølbak K, MiBa Board of Representatives. The Danish Microbiology Database (MiBa) 2010 to 2013. Euro Surveill 2014;19(1):20667.

4. Schmidt M, Schmidt SAJ, Sandegaard JL, Ehrenstein V, Pedersen L, Sørensen HT. The Danish National Patient Registry: a review of content, data quality, and research potential. Clinical Epidemiology 2015;449.
